# Neuroanatomical pathways of TMS therapy for depression

**DOI:** 10.1101/2025.02.10.25322034

**Authors:** Caio Seguin, L. Sina Mansour, Richard F. Betzel, Evgeny J. Chumin, Maria Grazia Puxeddu, Jinglei Lv, Linden Parkes, Luca Cocchi, Paul B. Fitzgerald, Robin F. H. Cash, Andrew Zalesky

**Author notes:** Authors contributed equally (Robin F. H. Cash, Andrew Zalesky).

## Abstract

We use connectome modelling to map polysynaptic fibre pathways underlying TMS therapy for depression. We propose putative cortical and subcortical routes connecting stimulation sites in the dorsolateral prefrontal cortex to the subgenual cingulate cortex via intermediate regions, and show route length explains (i) TMS treatment response in two independent patient cohorts and (ii) the clinical efficacy of fMRI-guided TMS targeting. Our results illuminate the neuroanatomical basis of TMS therapy for depression.

## Main text

Transcranial magnetic stimulation (TMS) of the left dorsolateral prefrontal cortex (DLPFC) is a therapy for refractory depression [1]. The therapy’s efficacy is thought to relate to indirect modulation of the subgenual cingulate cortex (SGC)—a structure that is not directly accessible to TMS. Support for this comes from observations that stimulating the DLPFC elicits acute and lasting changes in SGC activity and connectivity, as demonstrated by TMS combined with functional magnetic resonance imaging (fMRI) [2–4] and intracranial recordings [5]. Moreover, multiple fMRI studies indicate that baseline functional connectivity (FC) between DLPFC TMS sites and the SGC predicts patient treatment responses [6–9].

Interestingly, there is no evidence for strong axonal projections from the DLPFC to the SGC. Human brain imaging has not identified white matter tracts that could establish a direct communication channel between the two regions [10]. Tract-tracing work in rhesus monkeys has reported only sparse monosynaptic projections from the DLPFC to the SGC [11,12], instead arguing for an indirect route via pregenual area 32 [13]. The propagation of DLPFC stimulation to the SGC is therefore hypothesized to be polysynaptic, i.e., mediated by a sequence of intermediate regions interconnected via white matter fibres. Current methods to map white matter circuits using brain stimulation are limited to direct, “one-hop” fibres between anatomically connected regions [14,15], making them unsuitable for characterising DLPFC-SGC communication [16]. As a result, despite the progress achieved using fMRI, the neuroanatomical circuits mediating the therapeutic outcomes of TMS in depression remain elusive. This knowledge gap poses a central barrier to understanding and leveraging the role of the human structural connectome in mediating the response to TMS therapy.

Here, we use advances in connectome modelling to map polysynaptic fibre pathways from the DLPFC to the SGC and show that they explain TMS antidepressant effects. Network communication models quantify the capacity of signal transmission between regions via the connectome (Fig 1A) [17,18]. The models can identify multi-hop pathways (sequences of two or more white matter tracts) between grey matter structures that are not connected by a direct fibre. We use this modelling framework to reveal putative neuroanatomical routes traversed by TMS in two depression cohorts.

**Figure 1.**
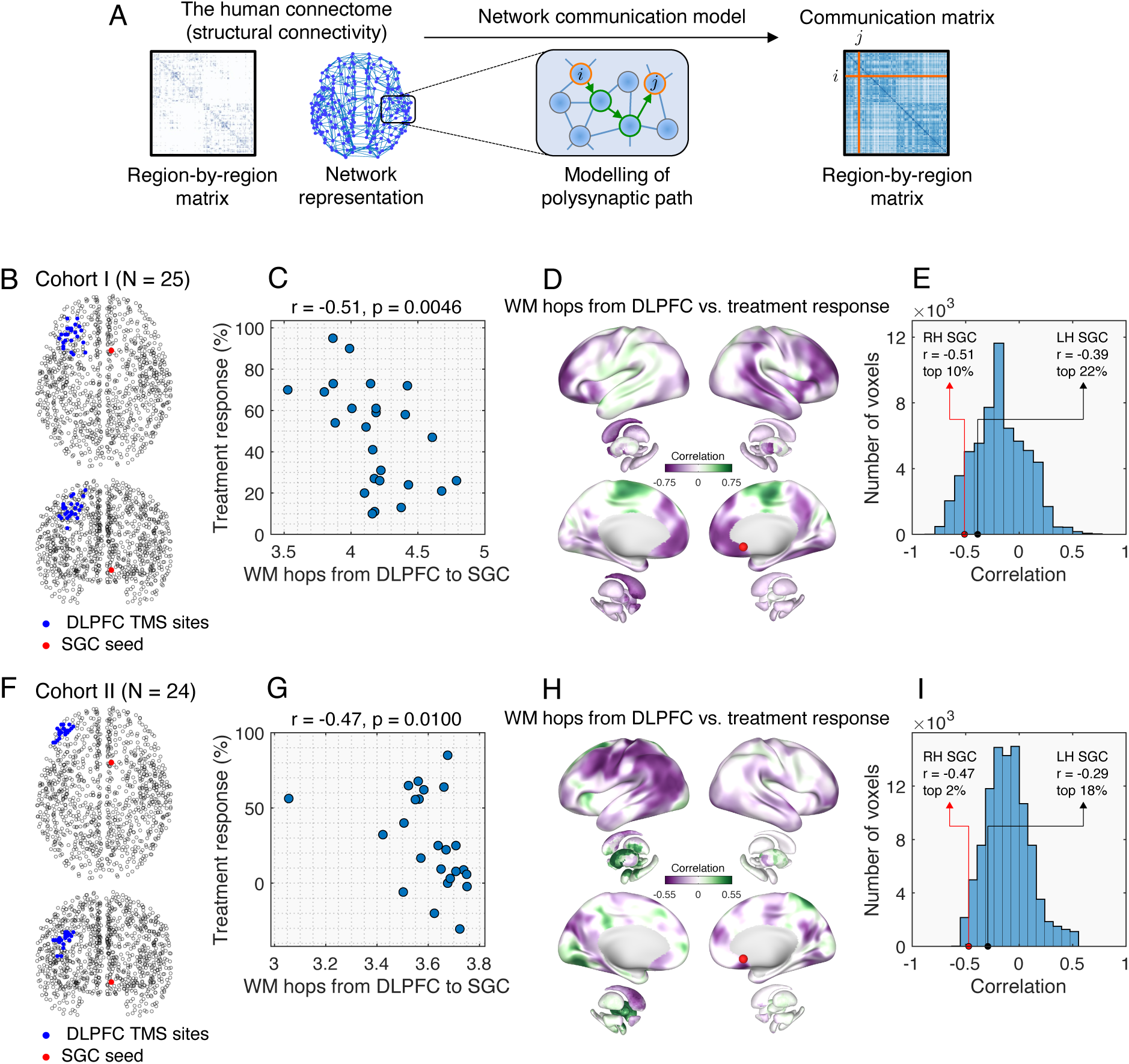
Association between DLPFC-SGC white matter pathways and TMS treatment response. **(A)** Network communication models quantify the capacity for signalling via the structural connectome. The models identify putative paths of polysynaptic communication between regions not connected by a direct fibre. **(B)** TMS and SGC coordinates in Cohort I. Black circles denote the grey matter regions of the connectome. **(C)** Relationship between treatment response and the number of white matter tracts traversed (WM hops) from DLPFC stimulation sites to the SGC, in Cohort I. Patients stimulated at DLPFC sites with shorter polysynaptic paths to the SGC showed better treatment outcomes. **(D)** Cortical map of the correlation between WM hops from DLPFC stimulation sites and treatment response, in Cohort I. In purple (green) are downstream regions for which efficient communication from the DLPFC was associated with better (worse) clinical outcomes. Treatment response was prominently associated with fewer WM hops from DLPFC stimulation site to right SGC (red dot, MNI [6,16, –10]), while also highlighting other regions of potential interest. **(E)** Distribution of correlations shown in (D). Voxels within TMS activation areas were excluded from the histogram. **(F-I)** Same as B-E but in Cohort II.

We consider two independent cohorts of individuals with treatment resistant depression who underwent repetitive TMS to the left DLPFC (Cohort I: *N* = 25, 17 females, 54.8 ± 9.9 years old [7]; Cohort II: *N* = 24, 10 females, 44.5 ± 11.8 years old [8]; *Materials and Methods*). Stimulation sites in the DLPFC were determined separately for each patient using established methods (Fig 1B,F). Treatment response was measured as the percentage change in the Montgomery–Åsberg Depression Rating Scale (cohort I) and Beck Depression Inventory (cohort II) scores before and after TMS therapy. We used network communication models to map polysynaptic fibre pathways from volumes of activated tissue centred at TMS coordinates in the left DLPFC to the right SGC (10-mm radius sphere centred at MNI [6,16, –10], as per [6–8]). We evaluated nine established network measures spanning different conceptualisations of how signals propagate through the connectome. We focus on pathways mapped using the *shortest path routing* model, which showed the most consistent associations across patient cohorts. In this model, a pathway’s capacity to support TMS propagation was measured as its average number of hops through the connectome (i.e., the number of white matter tracts traversed from the stimulation site to the SGC). Pathways were mapped on normative connectomes, constructed with the Lausanne (discovery) and Schaefer (replication) parcellations (*Materials and Methods*).

We found that polysynaptic white matter pathways were associated with treatment response in both cohorts. Patients stimulated at DLPFC sites connected to the SGC by fewer white matter hops—shorter, more direct communication channels—showed higher rates of clinical improvement (Lausanne connectome, cohort I: Spearman rank correlation coefficient *r* = – 0.51, one-tailed *p* = 0.0046, Fig 1C; cohort II: *r* = –0.47, *p* = 0.0100, Fig 1G). This suggests that white matter connectivity plays a key role in polysynaptic signalling from precise DLPFC stimulation sites to the SGC, and that variations in TMS coordinates—leading to differences in how efficiently signals are transmitted via the connectome—contribute to heterogeneity in patient outcomes.

To assess the robustness of this finding, we recomputed our analysis using an alternative connectome-mapping pipeline, comprising different image acquisition and pre-processing sequences, tractography algorithm, and grey matter parcellation. Once again, we found that patients stimulated at DLPFC sites with shorter polysynaptic paths to the SGC showed better treatment outcomes (Fig S1; Schaefer connectome, cohort I: *r* = –0.54, *p* = 0.0026; cohort II: *r* = –0.41, *p* = 0.0223). These associations were robust to leave-one-out outlier sensitivity tests (Fig S2). Out of the network measures evaluated, the number of steps along the shortest path was the only one associated with treatment outcome across both patient cohort and connectome-mapping pipelines (Fig S3; correlations were significant in 2 of out 4 scenarios following correction for multiple comparisons across nine network measures. Lausanne connectome: FDR Benjamini-Hochberg adjusted *p* = 0.0205; cohort II: *p* = 0.0897; cohort I, Schaefer connectome: *p* = 0.0205; cohort II: *p* = 0.20).

To further investigate this, we constructed surrogate connectomes by rewiring connections between brain regions. Surrogate connectomes are brain-like networks that preserve key features of white matter connectivity (e.g., the number of tracts intersecting each region, tract length distribution, and the relationship between tract weight and length) but are otherwise randomly shuffled into an alternative realisation of brain wiring [19,20]. We observed a significant decrease in the association between fibre communication and treatment response in surrogate connectomes (Fig S4), underscoring the importance of the biological topography of human white matter fibres to TMS propagation and its therapeutic efficacy in depression.

While there is accumulating evidence implicating the right SGC in TMS therapy for depression [2–9], could modulation of other downstream regions be more relevant to treatment response? To investigate this, we computed white matter pathways not only to the right SGC, but to 10-mm spheres centred at each grey matter voxel in the brain, yielding a whole-brain map of the correlation between TMS propagation and treatment response (Fig 1D,H). Our analysis corroborates clinical reports on the importance of the SGC seed centred at MNI [6,16, –10] [6–9], with this coordinate positioned at the top 10.17% and 2.25% of voxels most strongly associated with patient outcomes in cohorts I and II (Fig 1E,I, Lausanne connectome; Fig S1C,F, Schaefer connectome). Other prominent regions included portions of the medial superior, orbital frontal and superior temporal cortices as well as the precuneus and caudate. Interestingly, the contralateral left SGC seed (MNI [6,16,10]) yielded weaker correlations to treatment response, suggesting a lateralised role of this structure consistent with previous work [6,7]. We also note the presence of a few regions for which *longer* communication pathways were associated with better treatment response, suggesting that dampening TMS modulation of certain areas may also contribute to treatment efficacy.

Our analyses so far have presupposed a serial pathway of activation linking the DLPFC to the SGC. An alternative possibility is that TMS therapy is mediated by the parallel engagement of distributed downstream regions. For example, deep brain stimulation targets for depression often co-activate neighbouring structures within the cingulo-striatal circuit [21,22]. To test this, instead of the right SGC sphere, we repeated our analysis using a bilateral circuit mask encompassing the subgenual cingulate cortex, nucleus accumbens and caudate. Across both cohorts and connectome reconstructions, correlations with treatment response were comparable or stronger for the right SGC alone relative to the expanded circuit (Fig S5), supporting the specificity of the SGC as a key downstream target of DLPFC stimulation.

What neuroanatomical structures underpin TMS communication from the left DLPFC to the right SGC? To answer this question, we combined modelling results across individuals of the same cohort and counted the occurrences of the network paths used to infer white matter communication (Fig 2A,D). In agreement with evidence for a polysynaptic DLPFC-SGC route [10,13], we found that all paths were formed by a sequence of at least 2 steps, i.e., traversed at least one intermediate region (Fig 2B,E). Paths were consistent across both cohorts and mediated by key structures such as the thalamus, dorsal and medial areas of the superior frontal gyrus (SFG), and the anterior cingulate cortex (Fig 2C,F). The paths clustered into three main neuroanatomical routes—one cortical pathway and two complementary fronto-thalamic circuits (Fig 2H). The cortical route comprised three polysynaptic steps: from stimulation sites in the left DLPFC to the right dorsal SFG via the corpus callosum, to the right anterior cingulate cortex, to the right SGC via the cingulum bundle (Fig 2G; 2H, blue arrows). Both fronto-thalamic routes comprised four steps and differed primarily in their interhemispheric crossing point. The first and more frequently observed one proceeded from the left DLFC to the left thalamus, to the left medial SFG, crossed homotopically to the right SFG, and finally to the right SGC (Fig 2H, green arrows). The second fronto-thalamic route crossed hemispheres earlier, from the left DLPFC to the right dorsal SFG, to the right thalamus, to the right medial SFG, and to the right SGC (Fig 2H, orange arrows). We also note the presence of a minor ventral limbic pathway via the right thalamus, amygdala and temporal pole. Figures S6-S15 show the top five pathways from each cohort in MNI volumetric space.

**Figure 2.**
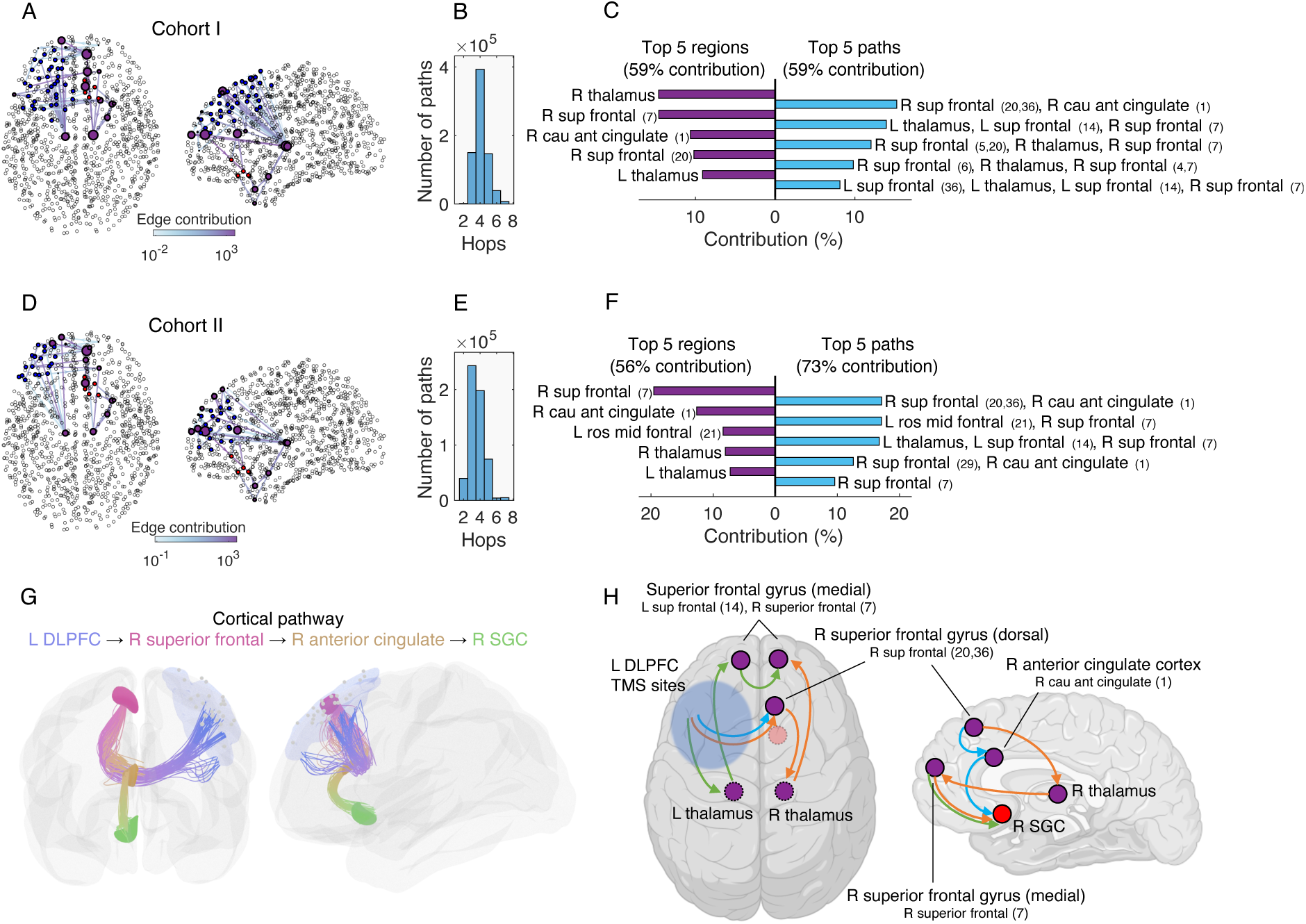
Neuroanatomical DLPFC-SGC pathways. **(A)** Network visualisation of paths originating from stimulated grey matter regions in the left DLPFC (blue), via intermediate regions (purple), to right SGC regions (red), in Cohort I. The size of intermediate regions and colour of connections are proportional to the number of times they are traversed. **(B)** Number of white matter hops from stimulated to SGC regions in Cohort I. **(C)** Top 5 intermediate regions and paths in Cohort I. Spatially adjacent grey matter parcels were grouped into the same parcellation labels for clarity (e.g., R sup frontal 20 and 36). **(D-F)** Same as (A-B) but in Cohort II. **(G)** Anatomical visualisations of the cortical pathway from left DLPFC to right SGC reconstructed using tractography. Fibre colours show different white matter tracts along the connectome. (H) Schematic of the three main neuroanatomical routes from the left DLFC to the right SGC. When relevant, regions are described with both anatomical nomenclature and labels from the Lausanne parcellation. The position of regions is approximate (see Figs S6-S15 for detailed MNI coordinates).

How do these neuroanatomical pathways advance on previous work on fMRI-guided TMS for depression? Prior evidence suggests that TMS is more effective when targeted to DLPFC sites with strong negative FC (i.e., anti-correlation) to the SGC [6–8] (although see [3]). As an initial check, we first confirmed this association in both depression cohorts using FC from the normative connectome datasets (Fig S3; Lausanne connectome, cohort I: *r* = –0.46, p = 0.0102, cohort II: *r* = –0.37, *p* = 0.0366; Schaefer connectome, cohort I: *r* = –0.48, p = 0.0079, cohort II: *r* = –0.35, p = 0.045). Although structural communication yielded stronger associations to treatment outcome than FC, this difference was not statistically significant (bootstrap non-parametric *p* > 0.05 for both cohorts and connectome datasets). Adding both predictors in a bivariate model did not improve explanatory power (all *F*-test *p* > 0.05), suggesting that structural communication and FC explain overlapping variance in treatment outcome.

Next, we computed two modalities of DLPFC maps: the first based on the FC to the SGC and second based on the number of hops via white matter pathways to the SGC (Fig 3). We found a positive correlation between the two maps, indicating that DLPFC sites with negative FC to the SGC also tend to be connected to it via shorter, more direct polysynaptic white matter pathways (Fig 3C,F; Lausanne connectome: *r* = 0.48; Schaefer connectome: *r* = 0.75; both *p* < 10^-12^). Lastly, we computed the DLPFC coordinates that minimise the polysynaptic path length to the SGC (Fig 3A,D; Lausanne connectome: MNI [–44,52,22], Schaefer connectome: MNI [–44,54,16]), and observed a negative correlation between the distance to these coordinates and clinical improvement, suggesting that stimulation nearer to this model-derived reference point tended to yield better outcomes (Fig S16). Together, these findings provide a neuroanatomical basis for the efficacy of fMRI-guided TMS targeting in depression and position connectome-based models as a promising framework for refining stimulation strategies.

**Figure 3.**
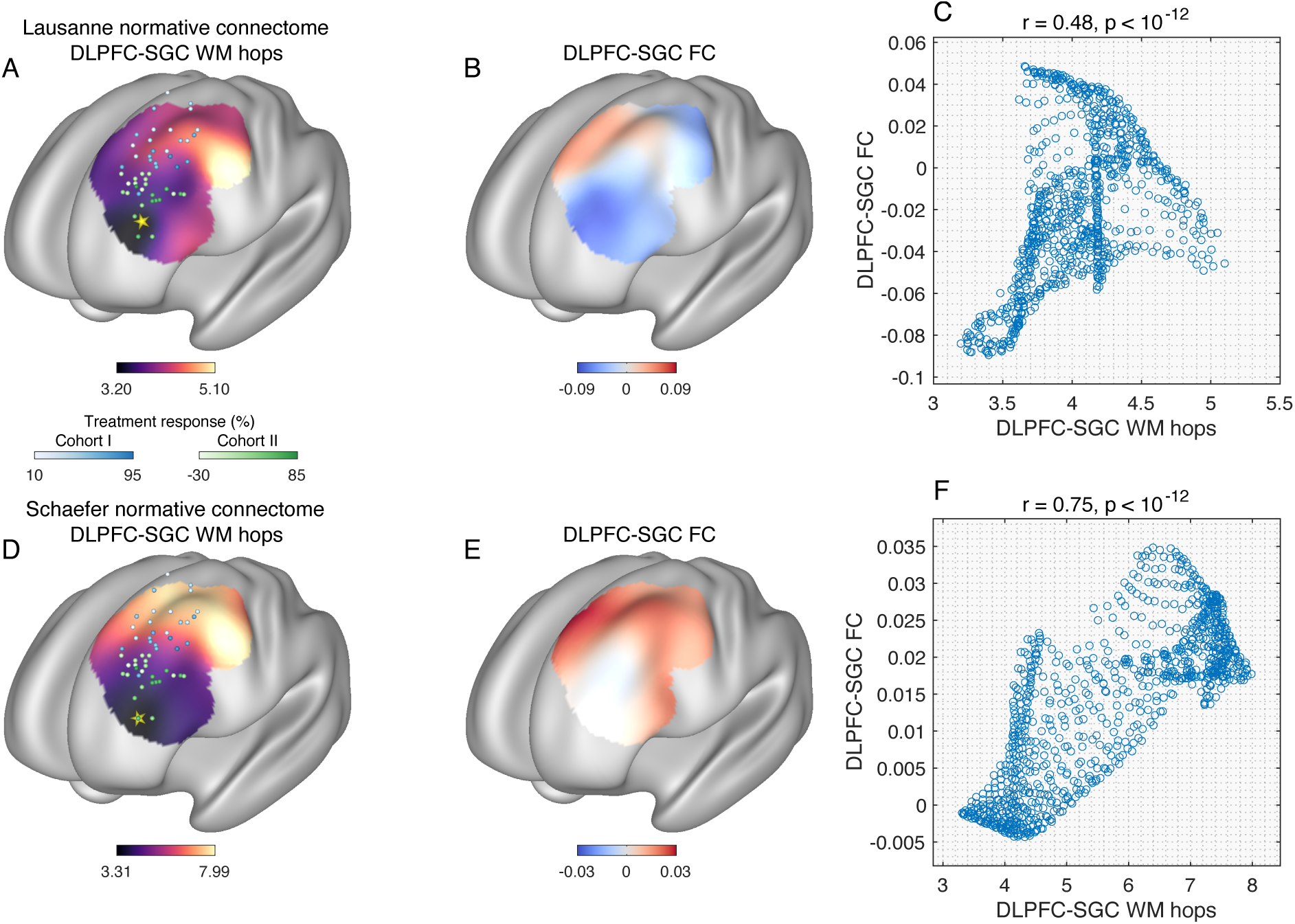
Structural and functional DLPFC-SGC maps. Maps show the number of white matter hops to the SGC **(A,D)** and the FC to the SGC **(B,E)** in two normative connectome datasets. Spheres mark TMS sites in cohorts I (blue) and II (green) and are coloured according to the patient’s treatment response. Gold stars mark the DLPFC loci that minimises the polysynaptic path length to the SGC (Lausanne: MNI [–44,52,22], Schaefer: MNI [– 44,54,16]). Scatter plots **(C,F)** show the association between DLPFC-SGC white matter communication and FC. Each dot represents a grey matter voxel in the DLPFC that is accessible to TMS. DLPFC sites functionally anti-correlated to the SGC tend to have shorter, more direct polysynaptic fibre pathways to the SGC.

In sum, we use network communication models to illuminate the neuroanatomical basis of DLPFC-targeted TMS for depression. We show that structurally-constrained communication from stimulation sites at the left DLPFC to the right SGC is correlated with patient outcomes. While the sample sizes in our analyses were modest, we replicate our results in two independent depression cohorts and two independent normative connectomes, demonstrating generalisability across different patient populations, TMS protocols, neuroimaging sequences, and connectome-mapping methods.

We propose cortical and subcortical circuits involved in therapeutic mechanisms of TMS for depression. The subcortical pathway supports literature implicating fronto-thalamic networks in depression [23–25], while the cortical pathway mediated by the anterior cingulate cortex (BA32) recapitulates tract-tracing findings in monkeys of a DLPFC-SGC route via pregenual area 32 [12,13]. Importantly, these pathways remain putative and are susceptible to the inherent limitations of diffusion MRI and tractography. For example, the cortical pathway progressed from the left DLPFC to the right medial superior frontal gyrus—a heterotopic connection that may reflect a tractography bias to terminate interhemispheric tracts along the medial wall [26]. Relatedly, cortico-thalamo-cortical circuits often involve relays via striatal and pallidal nuclei [27], but difficulties in tracking short range fibres within the basal ganglia may downplay the influence of these canonical loops in our models. Future work is therefore necessary to validate our findings, for example, by using intracranial recordings to track evoked responses along the proposed routes following DLPFC stimulation.

Our results establish the utility of structural connectivity—when coupled with computational models of neural communication [28–31]—to inform stimulation protocols based on polysynaptic fibre pathways. We focused on simple graph-theoretical models that estimate the capacity for inter-regional communication via the connectome. Their key advantages are interpretability and computational tractability, offering a straightforward method to quantify TMS propagation routes that can be readily translated into clinical interventions. In contrast, more complex biophysical models of neural dynamics may better capture the underlying neurophysiological mechanisms of TMS and will be valuable for future mechanistic work [30]. Among the network measures evaluated, the number of steps along the DLPFC-SGC shortest path was the strongest predictor of treatment response. This finding contrasts with earlier evidence that diffusion-based models more accurately predict electrical stimulation spread in the human connectome [28]. We speculate that diffusion processes may better reflect global patterns of network activation, whereas shortest paths could be more relevant for the targeted, circuit-specific effects mediating antidepressant response. Emerging datasets capturing real-time evoked activity from interleaved TMS-fMRI will help clarify these distinctions [4,32,33].

More broadly, a promising prospect is the use of computational models to augment current strategies for imaging-guided stimulation. Mapping polysynaptic paths may enhance existing methods based on direct tracts, such as fibre filtering and lesion network mapping [14,16]. We found that structural communication provides an anatomical grounding for fMRI-guided targeting in depression. Although neither imaging modality clearly outperformed the other in explaining treatment efficacy, prospective trials with larger sample sizes could explore integrating both in hybrid targeting strategies. In the future, our work could also be extended to evaluate and inform personalised targeting based on each individual’s unique connectome [9], across brain stimulation modalities including TMS and deep brain stimulation.

## Materials and Methods

### Clinical cohorts

We considered two previously published cohorts of individuals with treatment resistant depression who underwent repetitive TMS to the left DLPFC. Cohort I comprised 25 individuals (17 females, age 54.8 ± 9.9) treated at the Berenson-Allen Center at Beth Israel Deaconess Medical Center (BIDMC), Boston, USA [7]. The study was approved by the BIDMC’s Internal Review Board. Patients were diagnosed with major depression and met the criteria for medication-resistance, including failure to respond to at least one antidepressant. The treatment consisted of repetitive high-frequency TMS to the left DLPFC (10 and 20 Hz at 110% and 120% of the resting motor threshold, respectively) delivered daily for a period of 4-7 weeks. Precise stimulation sites were determined by moving 5.5 cm anterior to the motor threshold location along a parasagittal line. TMS coordinates were recorded using a neuronavigation system and later transformed to MNI space using the patients’ T1-weighted anatomical scans. Treatment response was measured by the change in the self-report of depressive symptoms using the Beck Depression Inventory (BDI-II), assessed on the first and last day of therapy. The mean baseline BDI score of 38.6 ± 9.3 was significantly decreased to 21.2 ± 13.0 following TMS treatment (*p* < 0.0001). Complete details on the clinical characterisation, data acquisition and TMS treatment for Cohort I are reported in [7].

Cohort II contained 24 individuals (10 females, 44.5 ± 11.8 years old) who completed 5-8 weeks of repetitive TMS to the left DLPFC as part of a larger clinical study (Australian New Zealand Clinical Trials Registry: Investigating Predictors of Response to Transcranial Magnetic Stimulation for the Treatment of Depression; ACTRN12610001071011) [8]. Individuals met the key inclusion criteria of major depression diagnosis and treatment resistant depression at Stage III of the Thase and Rush classification. Repetitive high-frequency TMS was delivered to the left DLPFC at 10 Hz and 110% of the motor threshold. Patient-specific stimulation coordinates were determined using the F3 beam approach and later transformed to MNI space using anatomical scans. Treatment response was measured as the difference in the self-report Montgomery–Åsberg Depression Rating Scale (MADRS) at baseline and 6 weeks after the start of TMS therapy. The mean baseline MADRS score of 33.7 ± 6.7 was significantly at 6 weeks to 24.3 ± 11.4 (*p* = 0.0003). Further details on the patient demographics, data acquisition and TMS treatment for Cohort II are found in [8] and https://www.anzctr.org.au/Trial/Registration/TrialReview.aspx?id=336262.

### Normative brain connectivity datasets

We used two previously published and openly available datasets of normative structural and functional connectivity. Normative connectomes—mapped from high-quality brain images of healthy adults—are commonly used to inform clinical brain stimulation studies as they have higher signal-to-noise ratio compared to connectivity reconstructed from patient data [14,34]. As detailed below, we used an established method to generate normative connectomes that mitigates tractography biases and filters out individual-level artefacts [35,36]. These factors contribute to the accurate delineation of white matter fibres and pathways associated with brain stimulation clinical outcomes [14,37].

The first normative dataset, used in all analyses in the main text, comprised brain networks for 70 healthy young adults (mean age 28.8 ± 9.1, 27 women) acquired at the University of Lausanne, Switzerland [38]. Grey matter was parcellated into 1,000 cortical and 14 subcortical (left and right thalamus, caudate, putamen, pallidum, accumbens area, hippocampus and amygdala) regions [39]. Structural connectivity was estimated from the diffusion MRI of individual participants using deterministic white matter tractography. The strength of the white matter connection between two regions was computed as the number of streamlines intersecting them divided by the average of their surface areas. Resting-state fMRI volumes underwent pre-processing to correct for motion, physiological confounds and global signal. The functional connectivity of each individual was computed as the Pearson correlation between regional time courses of blood oxygenation level dependent (BOLD) signals. Further details on image acquisition and connectivity reconstruction are found in [38].

The second dataset, used in the replication analysis of Fig S3, comprised brain networks from 1,000 Human Connectome Project (HCP) healthy young adults. Details on HCP imaging acquisition and pre-processing are described in [40–42]. Grey matter regions were defined based on the on the Schaefer 800 (7 Networks) parcellation [43] combined with the same 14 subcortical regions listed above. Structural connectivity was computed using probabilistic white matter tractography applied to diffusion MRI, and the strength of the structural connections was inferred as the streamline count between grey matter regions. Functional connectivity was once again computed as the Pearson correlation of regional BOLD time courses following resting-state fMRI pre-processing. Further details on connectivity reconstruction are provided in [44].

We applied an established consensus method to generate group-representative connectomes from the structural connectivity matrices of individual subjects [35]. The method selects the white matter connections most reliably found across individuals, while preserving the sample’s average connection density, numbers of intra- and inter-hemispheric connections, and distribution of connection lengths. This approach yields a prototypical map of human brain connectivity that mitigates tractography reconstruction errors, filters out individual-level artifacts, and boosts signal-to-noise ratio [35,36]. For the first dataset, this yielded a 1,014 × 1,014 structural connectivity matrix with 2.4% connection density. For the second dataset, we obtained a 814 × 814 structural connectivity matrix with 27.8% connection density.

### Neuroanatomically constrained connectomes

Structural connectivity matrices mapped using diffusion MRI and tractography are subject to well-established limitations. In particular, the presence of false-positive links can influence models of polysynaptic communication and lead to pathways that are not biologically realistic. For example, tract-tracing evidence in non-human primates shows that the subgenual cingulate cortex (SGC) sends monosynaptic projections to subcortical nuclei, but these links are unidirectional—oriented exclusively from the SGC to the subcortex. However, because diffusion MRI cannot resolve axonal directionality, tractography-based connections between the SGC and subcortical regions are bidirectional and encoded as symmetric entries in the structural connectivity matrices described above. The difficulty of reconstructing fibres with overlapping or complex geometries, such as those traversing the basal ganglia, can also generate spurious connections that are unlikely to exist in the human brain. While the connectomes considered in our study are publicly available and have been used in numerous studies [35,38,44], these limitations are particularly relevant to the goal of mapping polysynaptic communication pathways.

To address these issues, we pre-processed the structural connectivity matrices to constrain their connections using tract-tracing evidence from non-human primates. This procedure systematically and conservatively removed links that are likely to be false positives based on established neuroanatomical knowledge. Both normative connectomes considered in this study—the Lausanne (1,014 regions) and Schaefer (814 regions) parcellations—include the same 14 FreeSurfer subcortical regions (left and right thalamus, caudate, putamen, pallidum, accumbens, hippocampus, and amygdala). Connections to and from subcortical structures were the focus of the neuroanatomical constraints. The tract-tracing literature provides well-established knowledge on the anatomy of the primate subcortex, with clear homology between the human and monkey subcortical structures. This makes it possible to use tract-tracing evidence to refine the connections intersecting large-scale subcortical structures in human connectivity matrices (we did not consider subdivisions of regions into finer nuclei). In contrast, extending tract-tracing evidence to constrain thousands of cortico-cortical links would be intractable and less critical, as most cortico-cortical projections are bidirectional [45–47], reducing the risk of false positives due to the lack of directional information. Based on these points, the following neuroanatomical constraints were applied:

● **Cortex ↔ cortex:** All cortico-cortical connections were retained as defined in the original matrices, consistent with evidence that most cortical projections are bidirectional, although their strength may vary with direction [45–47].
● **Subcortex → cortex:** Projections to the cortex were retained only for the thalamus [48,49], hippocampus [50,51], and amygdala [52,53]. Outgoing links from striatal and pallidal regions (caudate, putamen, accumbens, and pallidum) to the cortex were excluded, as these projections are unidirectional and originate in the cortex [27,54,55].
● **Cortex → subcortex:** With the exception of the pallidum, all subcortical regions in our parcellation receive projections from the cortex, including the striatum [27,54,55], thalamus [48,49], hippocampus [50,51], and amygdala [52,53]. Projections from the cortex to subcortical structures were therefore retained, except those targeting the pallidum [56].
● **Subcortex ↔ subcortex:** Connectivity within the subcortex was constrained according to established tract-tracing data:

○ **Thalamus:** Sends unidirectional projections to the caudate, putamen and accumbens; receives unidirectional projections from the pallidum; and has bidirectional projections with the hippocampus and amygdala [48,49].
○ **Caudate and putamen:** Receive unidirectional projections from the thalamus; and have bidirectional projections to the pallidum (the FreeSurfer pallidum combines the internal and external globi pallidi) [27,54,55].
○ **Pallidum:** Sends unidirectional projections to the thalamus; receives unidirectional projections from the accumbens and amygdala; and exchanges bidirectional projections with the caudate and putamen [27,56].
○ **Accumbens:** Sends unidirectional projections to the pallidum and receives unidirectional projections from the thalamus, hippocampus and amygdala [27,54,55].
○ **Hippocampus:** Sends unidirectional projections to the accumbens; has bidirectional projections with the amygdala and thalamus; and has bidirectional homotopic projections between left and right hippocampi [50,51].
○ **Amygdala:** Sends unidirectional projections to the pallidum and accumbens; has bidirectional projections to the thalamus and hippocampus; and has bidirectional homotopic projections between left and right amygdalae [52,53].
● **Contralateral connections:** Cortico-subcortical and subcortical-subcortical links were restricted to ipsilateral projections, with the exception of the interhemispheric connections between left and right hippocampi and amygdalae listed above. This decision is supported by extensive evidence that subcortical circuitry is dominated by ipsilateral connectivity in primates [54,57,58]. While tracer studies have identified contralateral corticothalamic and cortico-striatal projections, these are sparser and vastly outnumbered by their ipsilateral counterparts [57,58].

The neuroanatomical constraints are summarised in the connectivity mask shown in Fig S17. A matrix entry *ij* indicates the presence (blue) or absence (white) of monosynaptic projections from structure *i* to structure *j*. This mask was applied to both normative connectomes via element-wise multiplication, thereby removing likely false positive links while preserving the tractography-derived weights of the remaining connections. For the Lausanne connectome, this process removed 1,882 links, yielding a directed connectivity matrix with 2.3% density (a reduction of 0.1% compared to the original 2.4%). For the Schaefer connectome, 8,699 links were removed, resulting in a directed connectivity matrix with 26.58% density (1.3% reduction compared to the original 27.89%). All analyses were carried out using the neuroanatomically constrained connectomes.

Finally, we note that while these constraints improve the biological validity of the connectomes, they rely on simplifying assumptions and may have drawbacks, such as potential anatomical differences between human and non-human primates. Therefore, for completeness, we replicated key analyses using unconstrained connectomes (Fig S18).

### Brain network communication models

In the main text, we computed brain network communication using the *shortest path routing*. This model identifies the shortest, most direct channel of communication between two brain regions. For a pair of regions that do not share a direct white matter connection—e.g., the DLPFC and the SGC—the shortest path comprises a sequence of two or more white matter connections via the connectome. We considered the weighted shortest path, such that the length of a path is quantified by taking into account the total strength of the fibres comprising it.

Formally, let *W* be the structural connectivity matrix, where entry *W*(*i*, *j*) denotes the strength of the white matter connections between regions *i* and *j*. We first define the matrix of connection lengths *L* = −*log* (*W*) to quantify the distance or travel cost between pairs of regions. The shortest path between two regions *a* and *b* is the sequence of connections Π(*a*, *b*) = {*a*, *u*, …, *v*, *b*} that minimises the sum *L*(*a*, *u*) + ⋯ + *L*(*v*, *b*), known as the shortest path length. The capacity for TMS communication from *a* to *b* was defined as the number of hops along their shortest path, such that *H*(*a*, *b*) = |Π(*a*, *b*)|. Computed for every pair of regions, this yielded the communication matrix *H* (Fig 1A).

In Figure S3, we explored alternative brain network communication measures that do not quantify communication via shortest paths. The definitions and technical details of these measures can be found in [17,28,59] and code for their computation is available in the *Brain Connectivity Toolbox* [60]. Importantly, each measure establishes a clear hypothesis on the direction of association with TMS propagation. For instance, the number of hops along the shortest path quantifies the difficulty or cost of communication, such that paths with more hops are expected to support weaker TMS transmission compared to paths with fewer hops. Given the clear hypotheses on the directions of associations, we considered one-tailed *p*-values when testing for correlations between network communication measures and treatment response.

### Modelling communication from DLPFC TMS sites to the SGC

We computed the average number of white matter hops via the shortest path from TMS sites to the 10-mm radius SGC sphere centred at MNI [6,16, –10]. We first registered TMS coordinates and grey matter parcellations to MNI space. In some cases, recorded stimulation coordinates were positioned slightly outside of the MNI standard brain template and were shifted to the closest within-template grey matter voxel based on Euclidean distance. TMS was modelled to activate grey matter voxels within a 10-mm radius of the stimulation coordinate. Similarly, the SGC sphere comprised grey matter voxels within a 10-mm radius of the SGC coordinate. To ensure analyses captured communication to the subgenual cingulate cortex, any subcortical voxels in the SGC sphere were discarded. Overlaid onto a grey matter parcellation, each voxel *i* in the stimulation and SGC spheres was mapped to a parcel *p*(*i*) corresponding to a row/column of the structural connectivity matrix.

Communication from patient *k*’s TMS site to the SGC was computed as a distance-weighted average of the number of hops between voxels in the stimulation and target spheres. Each voxel *i* in the stimulation sphere was assigned a weight *w*(*i*) = *e*^kd(i)^, *K* ≤ 0, where *d*(*i*) is the Euclidean distance of voxel *i* from the sphere’s centre, such that voxels nearer the TMS coordinate contributed more strongly. Voxels in the SGC sphere were uniformly weighted. Communication was then defined as

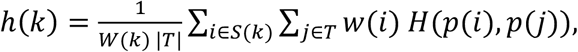

where *H*(*p*(*i*), *p*(*j*)) is the number of hops along the shortest path between the parcels of voxels *i* and *j*, and *W*(*k*) = Σ_i∈s)_*w*(*i*).

We note that *H* is computed on a group-representative connectome, and therefore variations in communication across patients are encoded in *S*(*k*) and stem solely from differences in TMS coordinates. In analyses where communication was modelled to loci other than the SGC, the composition of *T* adjusted accordingly (e.g. Fig 1D,H). In the analyses of Fig S3, *H* was replaced by communication matrices from alternative network measures and by functional connectivity matrices. For network measures, we set the decay parameter *κ* = –1, reflecting the spatial specificity of white matter pathways, where small variations in the stimulation site may engage different discrete fibres. For functional connectivity, consistent with previous work [6–8], we used a uniform weighting (*κ* = 0), which better reflects the spatial smoothness of fMRI data. Sensitivity analyses (Fig. S19) confirmed that results remained stable across a range of κ values. The communication pathways in Fig 2 were identified by summing the weighted contributions of paths connecting stimulation and target voxels across patients within each cohort.

### Visualisation

The cortical maps in Fig 1D,H were rendered using *Cerebro* [61]. To visualise the white matter pathways in Fig 2G, we first performed tractography on a tissue-unbiased template image of fibre orientation distributions created using multimodal registration [62]. The template was generated in previous work using multimodal registration to guarantee the alignment of white matter tracts with cortical and subcortical grey matter [62]. Streamlines were then transformed to MNI space and visualised using *Cerebro* [61] and *Blender*. All other figures were created using *MATLAB*.

## Code availability

Code to reproduce the analyses of the present manuscript is available at https://github.com/caioseguin/tms_dep_pathways. Network communication models were computed using the *Brain Connectivity Toolbox* [60]. Surrogate connectomes were computed using code available through reference [20].

## Data Availability

Clinical data (TMS coordinates and rates of symptom improvement) from Cohort I is publicly available as part of the Supplementary Information of reference [7]. Clinical data from Cohort II (ACTRN12610001071011) [8] remain subject to privacy and ethical restrictions. The two normative connectome datasets are publicly available through references [38,44]. The publicly available data used in the analyses of the present manuscript can be found at https://github.com/caioseguin/tms_dep_pathways.

## Supplementary Information

**Figure S1.**
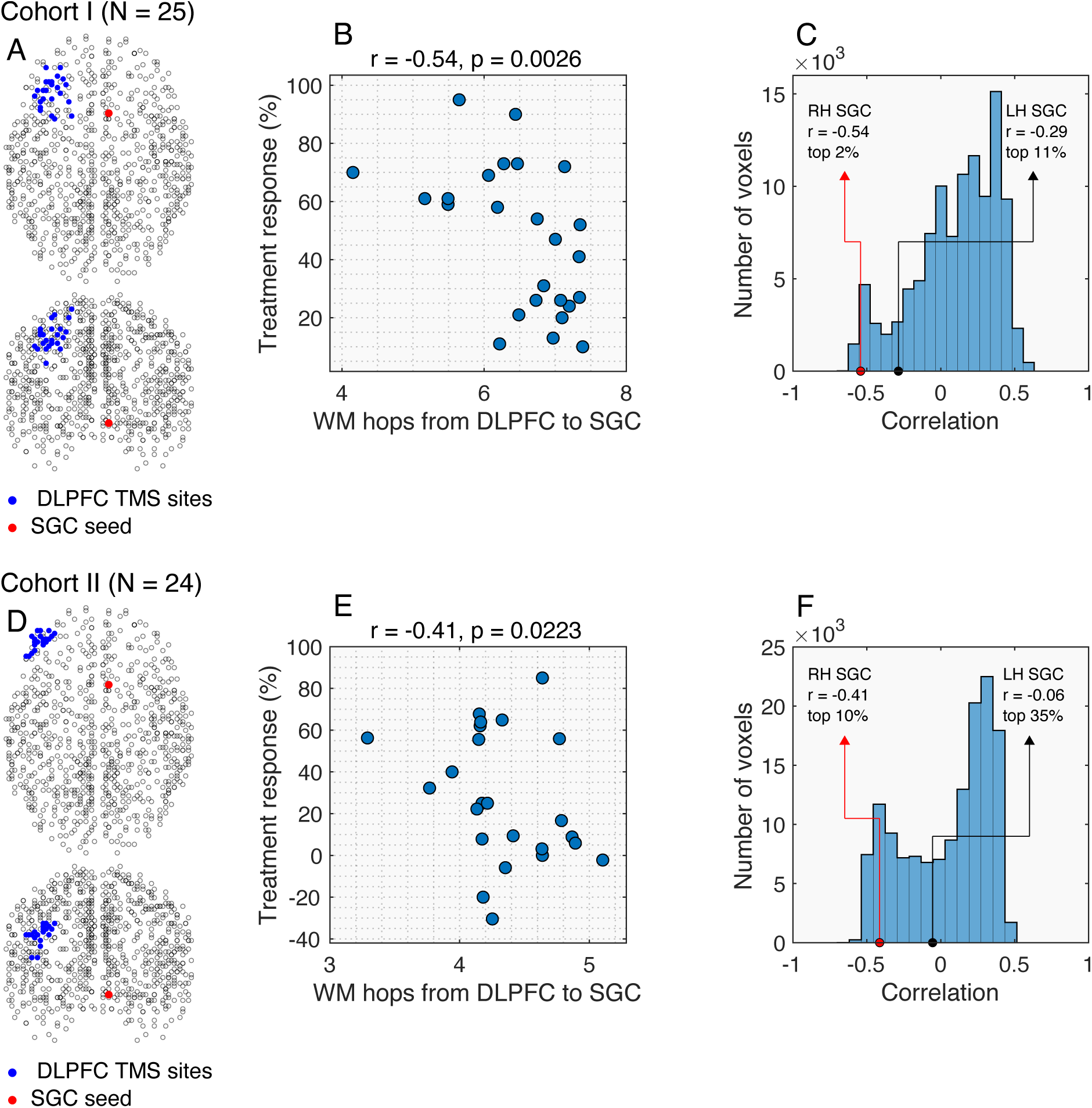
Association between DLPFC-SGC white matter pathways and TMS treatment response in the Schaefer normative connectome. **(A)** TMS and SGC coordinates in Cohort I. Black circles denote the grey matter regions of the connectome. **(B)** Relationship between treatment response and the number of white matter tracts traversed (WM hops) from DLPFC stimulation sites to the SGC, in Cohort I. Patients stimulated at DLPFC sites with shorter polysynaptic paths to the SGC showed better treatment outcomes. **(C)** Distribution of correlations between treatment response and WM hops from DLPFC stimulation sites to all grey matter voxels. Voxels within TMS activation areas were excluded from the histogram, in Cohort I. **(D-F)** Same as B-E but in Cohort II.

**Figure S2.**
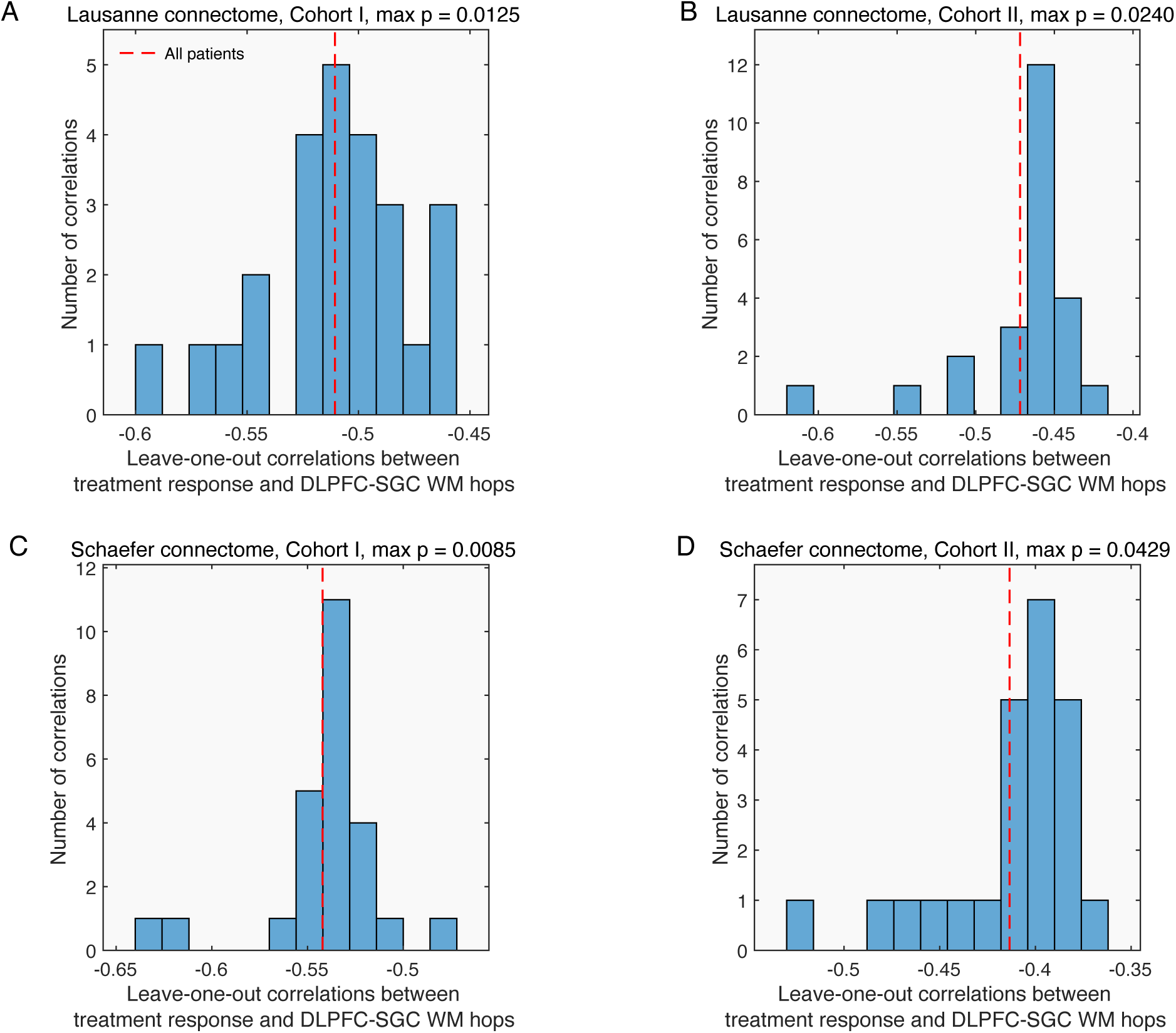
Leave-one-out sensitivity analyses. Correlations between DLPFC-SGC white matter hops and treatment response were computed by leaving out one patient at a time. Across both patient cohorts and normative connectomes, all leave-one-out correlations were significant (Lausanne connectome, Cohort I: Spearman correlation maximum *p* = 0.0125; cohort II: *p* = 0.0240; Schaefer connectome, Cohort I: *p* = 0.0085; Cohort II: *p* = 0.0429), indicating that the associations are not driven by the inclusion or exclusion of any single patient. Dashed red lines show the correlations obtained considering all patients, as reported in the Main Text.

**Figure S3.**
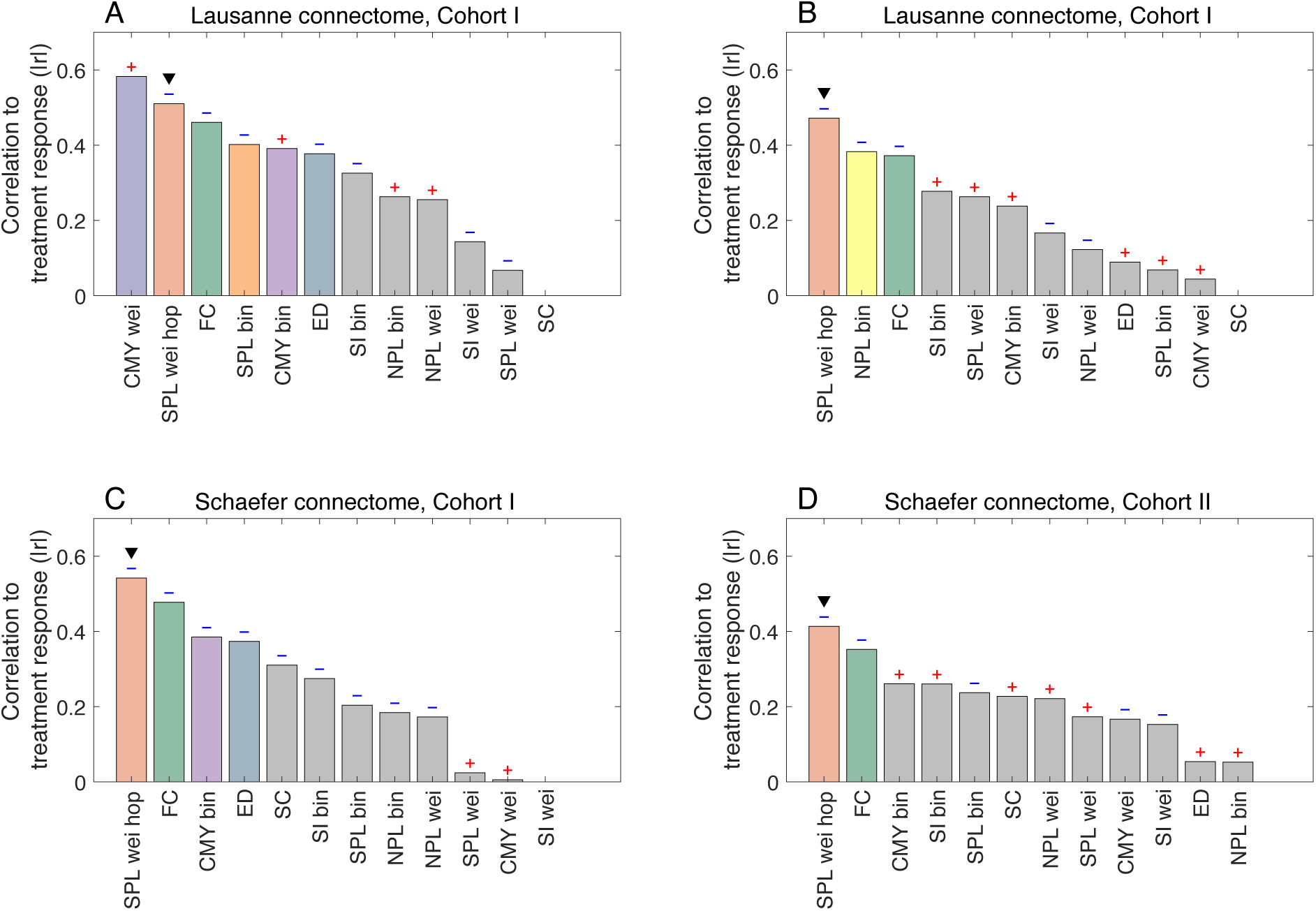
Correlations between treatment response and various connectivity, communication and distance measures. Correlations were computed in two cohorts of patients suffering from depression who received TMS to the left DLPFC and using two normative connectome datasets. All measures capture interactions between the DLPFC stimulation sites and the SGC, and were computed following the procedures described in Methods sections *Neuroanatomically constrained connectomes*, *Brain network communication models*, and *Modelling communication from DLPFC TMS sites to the SGC*. Each coloured bar shows the absolute value of the Spearman correlation coefficient between treatment response and one measure, with signs atop bars indicating the direction of association. Measures include functional connectivity (FC), structural connectivity (SC), Euclidean distance (ED), and nine network communication measures computed on binary (bin) and weighted (wei) connectomes. Briefly, the network communication measures are the number of hops of the weighted shortest path (SPL wei hop, used in the main text analyses and highlighted by triangular markers), the binary and weighted shortest path lengths (SPL bin, wei), the binary and weighted navigation path lengths (NPL wei, bin), the binary and weighted search information (SI wei, bin), and the binary and weighted communicability (CMY wei, bin). Technical details on these measures are described in [17,28,59]. Bars in grey denote correlations that were not significant (Spearman rank correlation one-tailed *p* > 0.05). The number of hops of the weighted shortest path (SPL wei hop) was the only model with significant correlations across the four combinations of depression cohorts and normative connectome datasets (*p* = 0.0046, 0.0100, 0.0026, 0.0223, for panels A to D, respectively). In two of out of four dataset combinations (panels A and C), these associations remained significant after correcting for multiple comparisons across the nine network communication measures using the FDR Benjamini-Hochberg method (adjusted *p* = 0.0205, 0.0897, 0.0205, 0.20 for panels A to D, respectively).

**Figure S4.**
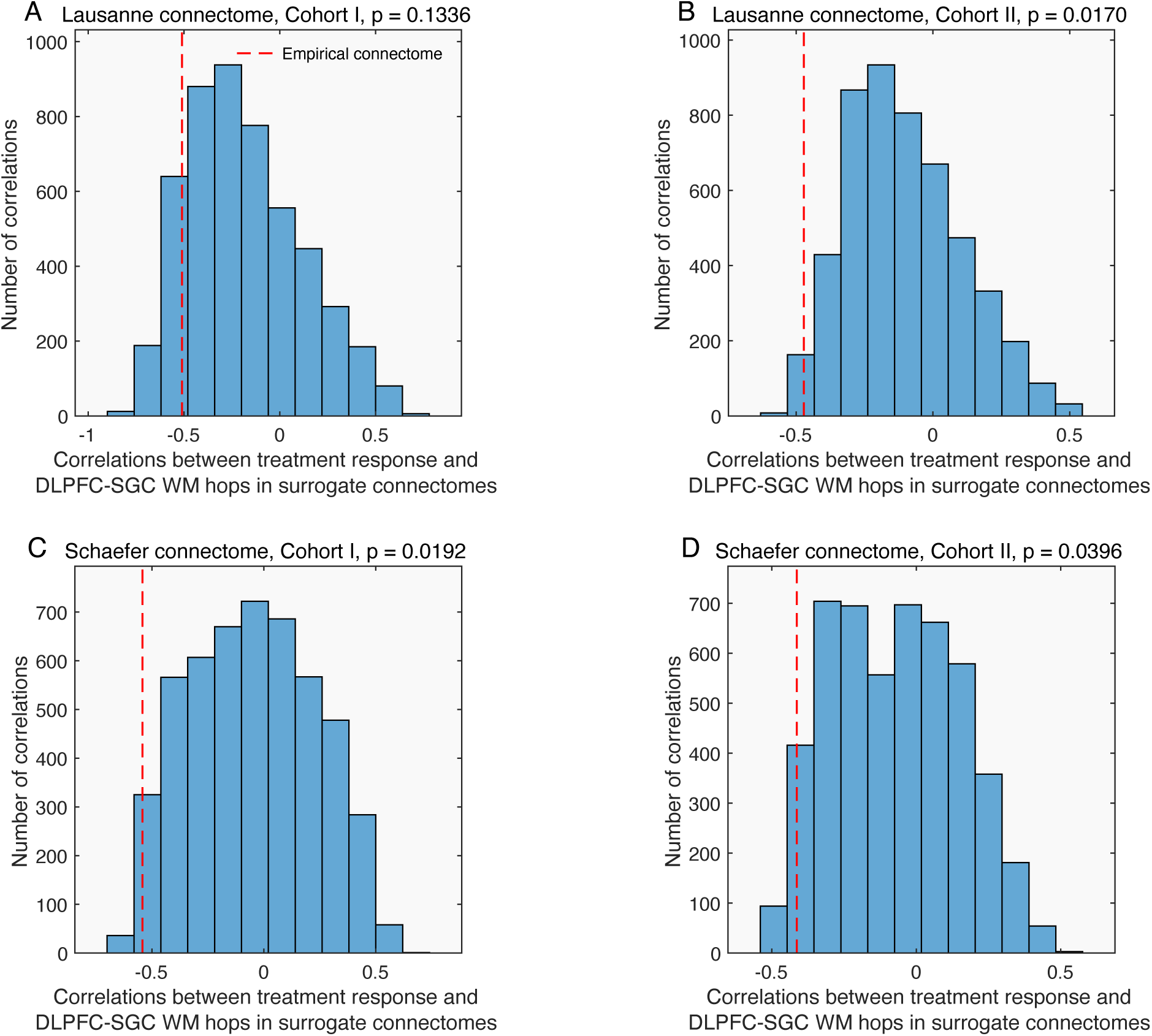
Surrogate connectome analyses. Dashed red lines show the correlations obtained considering empirical connectomes, as reported in the Main Text. We constructed surrogate connectomes by rewiring connections between brain regions. Surrogate connectomes are brain-like networks that preserve key features of structural connectivity (e.g., the number of tracts intersecting each region, tract length distribution, and the relationship between tract weight and length) but are otherwise randomly shuffled into an alternative realisation of brain wiring [19,20]. We repeated our analyses in an ensemble of 5,000 surrogate connectomes. Across patient cohorts and normative connectomes, we observed a marked decrease in the association between white matter communication and treatment response, which was statistically significant in three out of four dataset combinations (Lausanne connectome, Cohort I: non-parametric *p* = 0.1336; Cohort II: *p* = 0.0170; Schaefer connectome, Cohort I: *p* = 0.0192; Cohort II: *p* = 0.0396). This underscores the importance of the detailed topography of human white matter fibres to TMS propagation and its therapeutic efficacy in depression.

**Figure S5.**
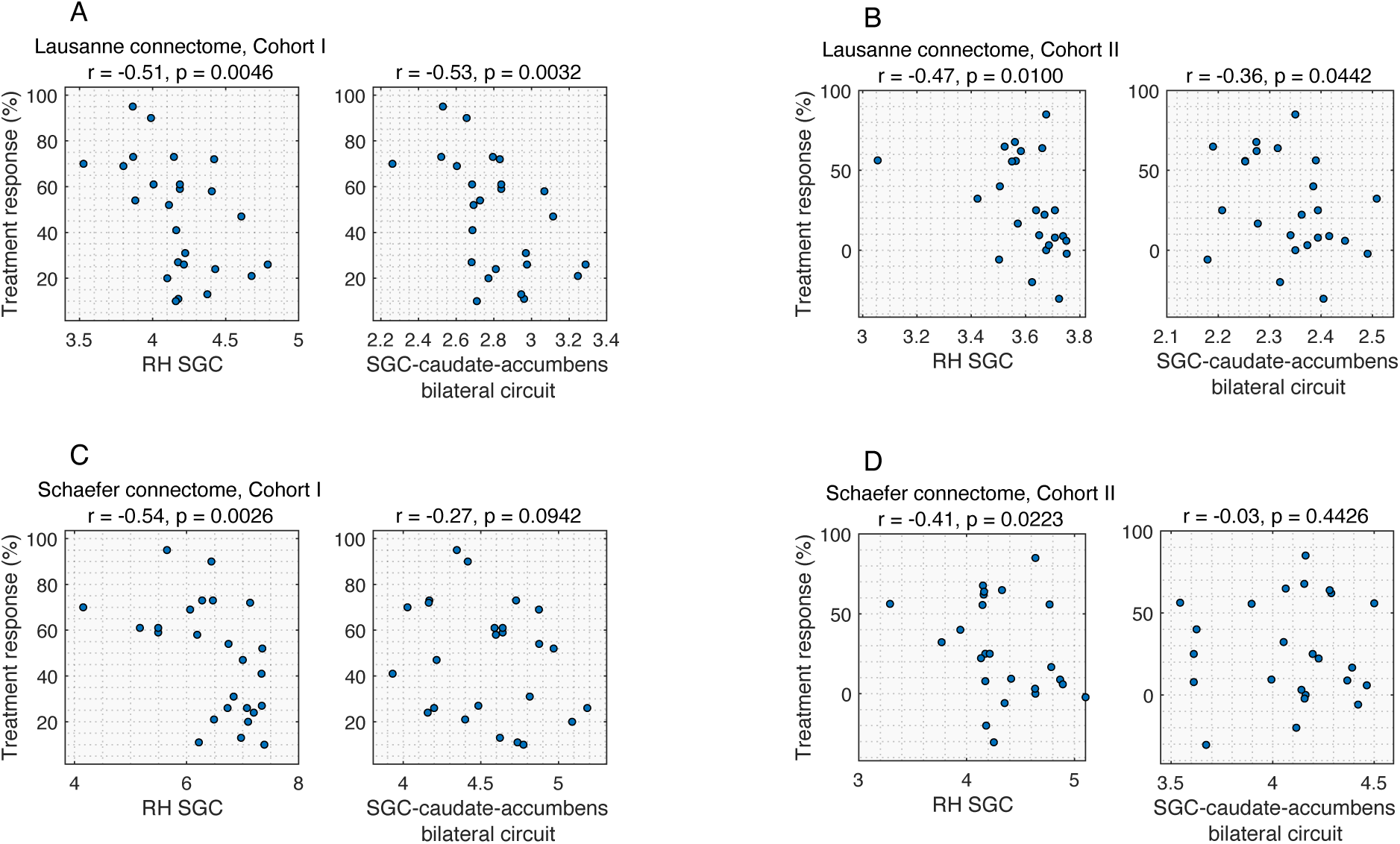
Parallel cingulo-striatal circuit analyses. **(A)** Lausanne connectome, Cohort I. Left: Associations between treatment response and communication from DLPFC TMS sites to the SGC, as reported in the Main Text (Fig 1C). Right: Associations considering communication from DLPFC TMS sites to a parallel cingulo-stratial circuit including the right and left SGC, caudate and accumbens area. Other panels show the same analyses for **(B)** Lausanne connectome, Cohort II, **(C)** Schaefer connectome, Cohort I, and **(D)** Schaefer connectome, Cohort II. Across both cohorts and connectome reconstructions, correlations with treatment response were comparable or stronger for the right SGC alone relative to the expanded circuit, supporting the specificity of the SGC as a key downstream target of DLPFC stimulation.

**Figure S6.**
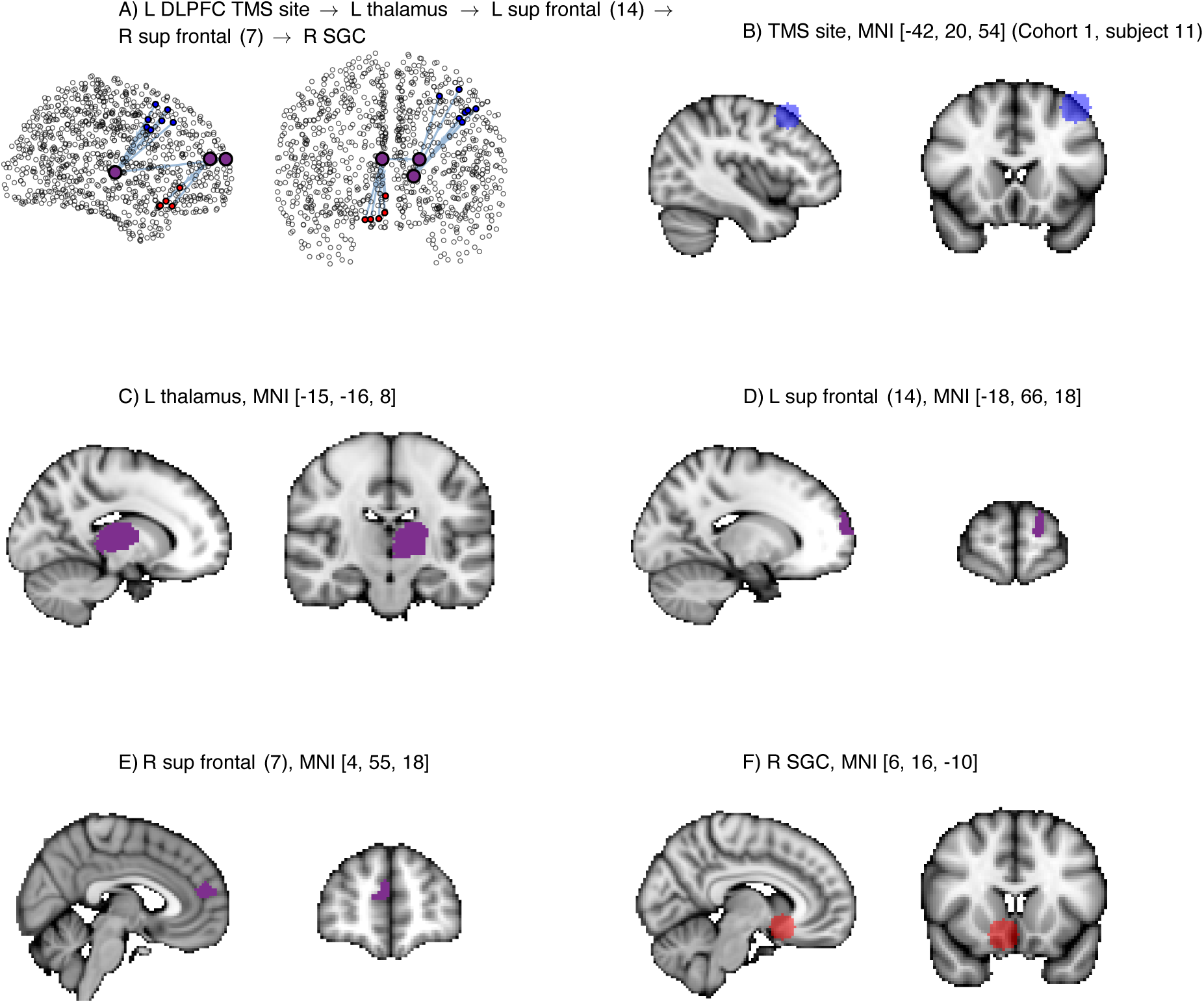
DLPFC-SGC path #1, Cohort I, visualised in network format and volumetric MNI-space for an exemplary patient. Grey matter regions along the path are identified by their Lausanne parcellation label (scale 5) and centroid MNI coordinate.

**Figure S7.**
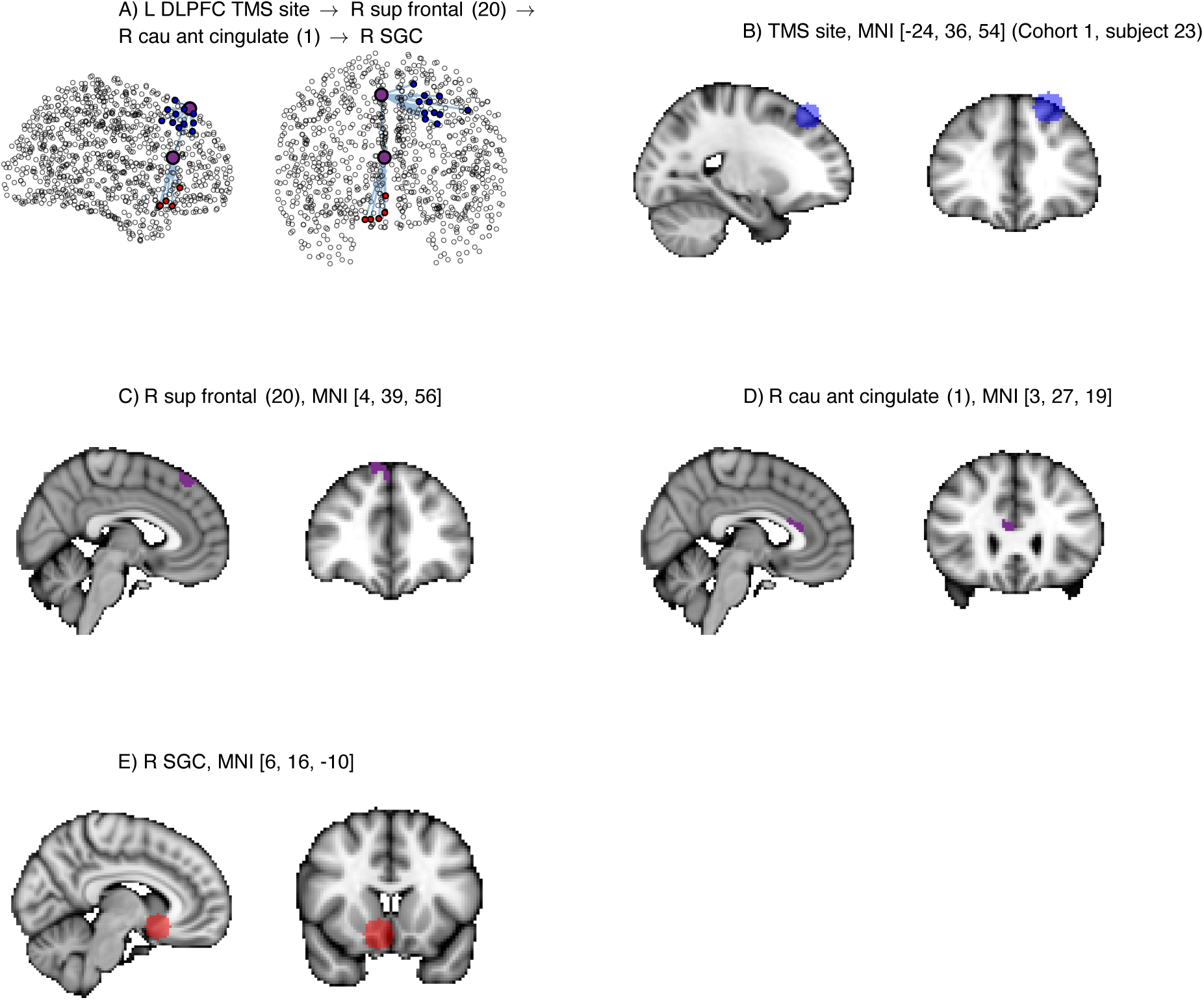
DLPFC-SGC path #2, Cohort I, visualised in network format and volumetric MNI-space for an exemplary subject. Grey matter regions along the path are identified by their Lausanne parcellation label (scale 5) and patient MNI coordinate.

**Figure S8.**
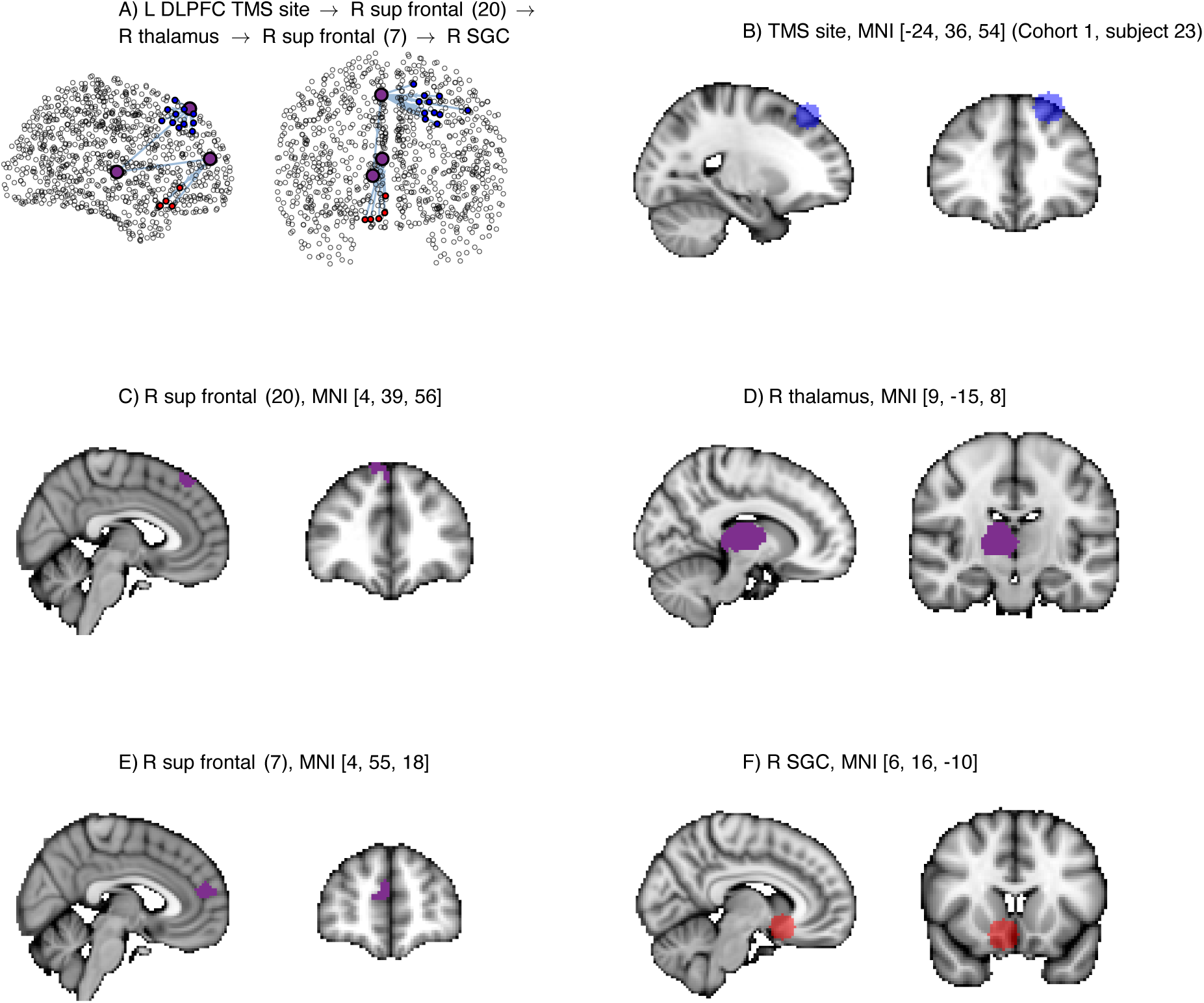
DLPFC-SGC path #3, Cohort I, visualised in network format and volumetric MNI-space for an exemplary patient. Grey matter regions along the path are identified by their Lausanne parcellation label (scale 5) and centroid MNI coordinate.

**Figure S9.**
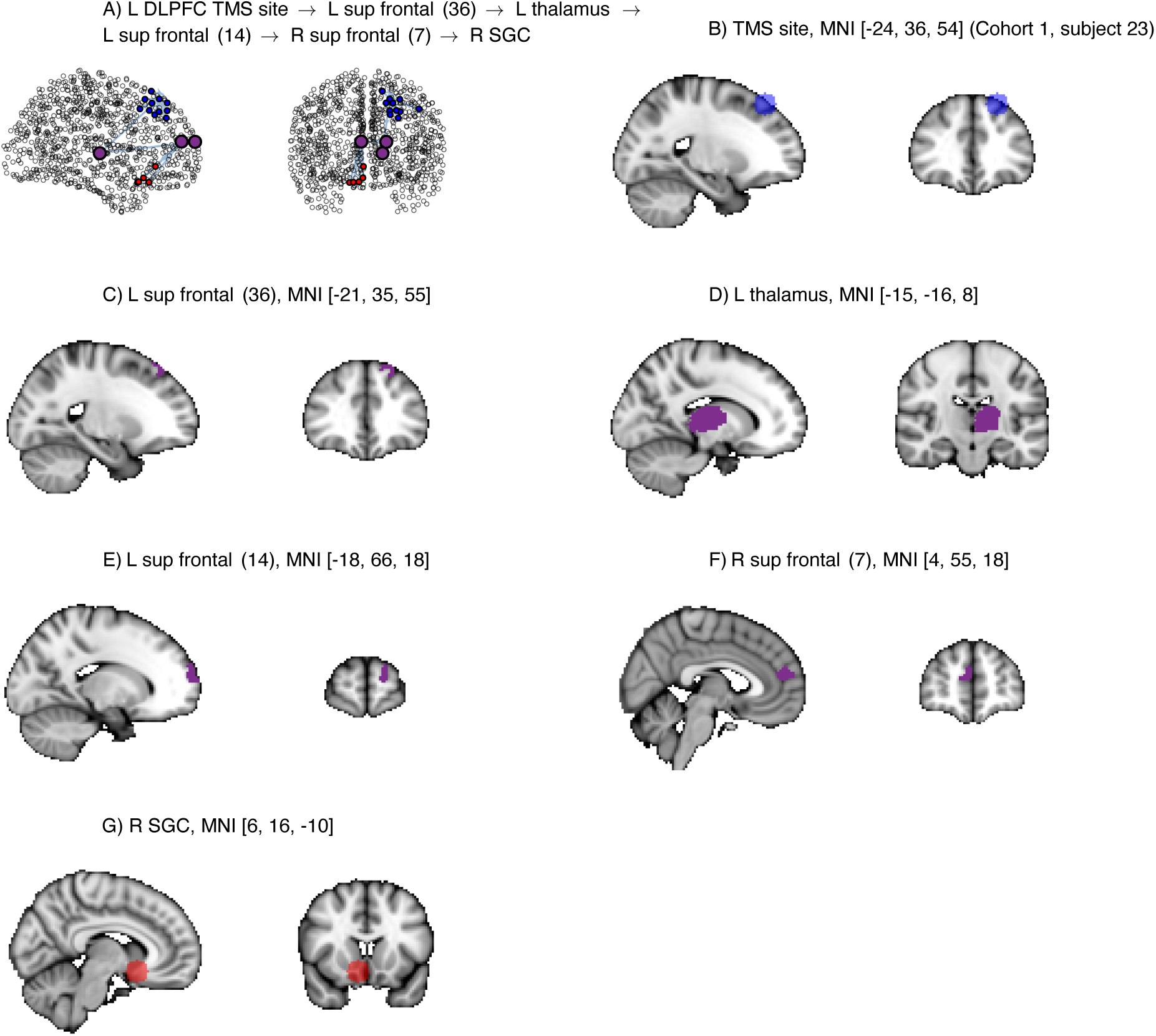
DLPFC-SGC path #4, Cohort I, visualised in network format and volumetric MNI-space for an exemplary patient. Grey matter regions along the path are identified by their Lausanne parcellation label (scale 5) and centroid MNI coordinate.

**Figure S10.**
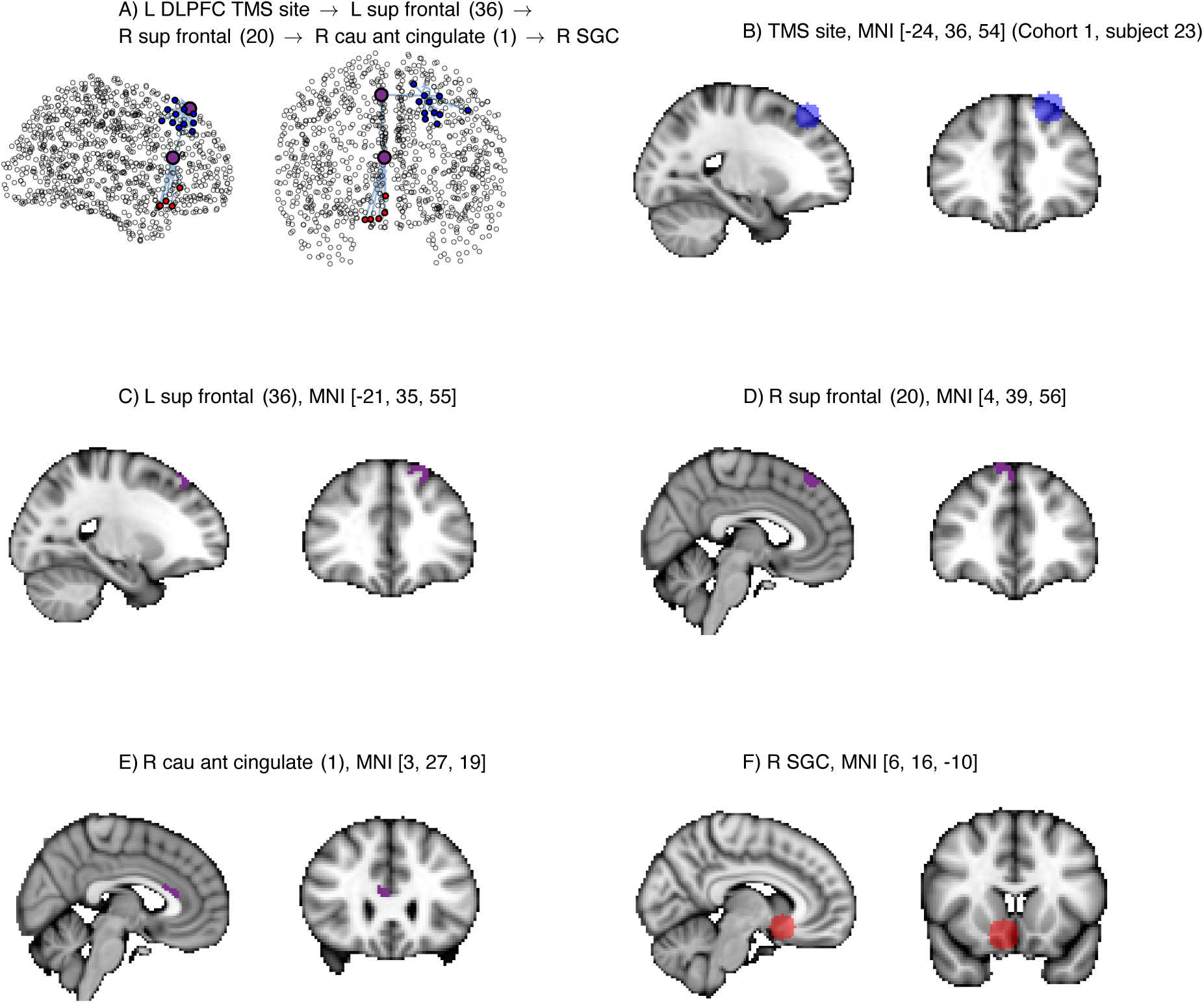
DLPFC-SGC path #5, Cohort I, visualised in network format and volumetric MNI-space for an exemplary patient. Grey matter regions along the path are identified by their Lausanne parcellation label (scale 5) and centroid MNI coordinate.

**Figure S11.**
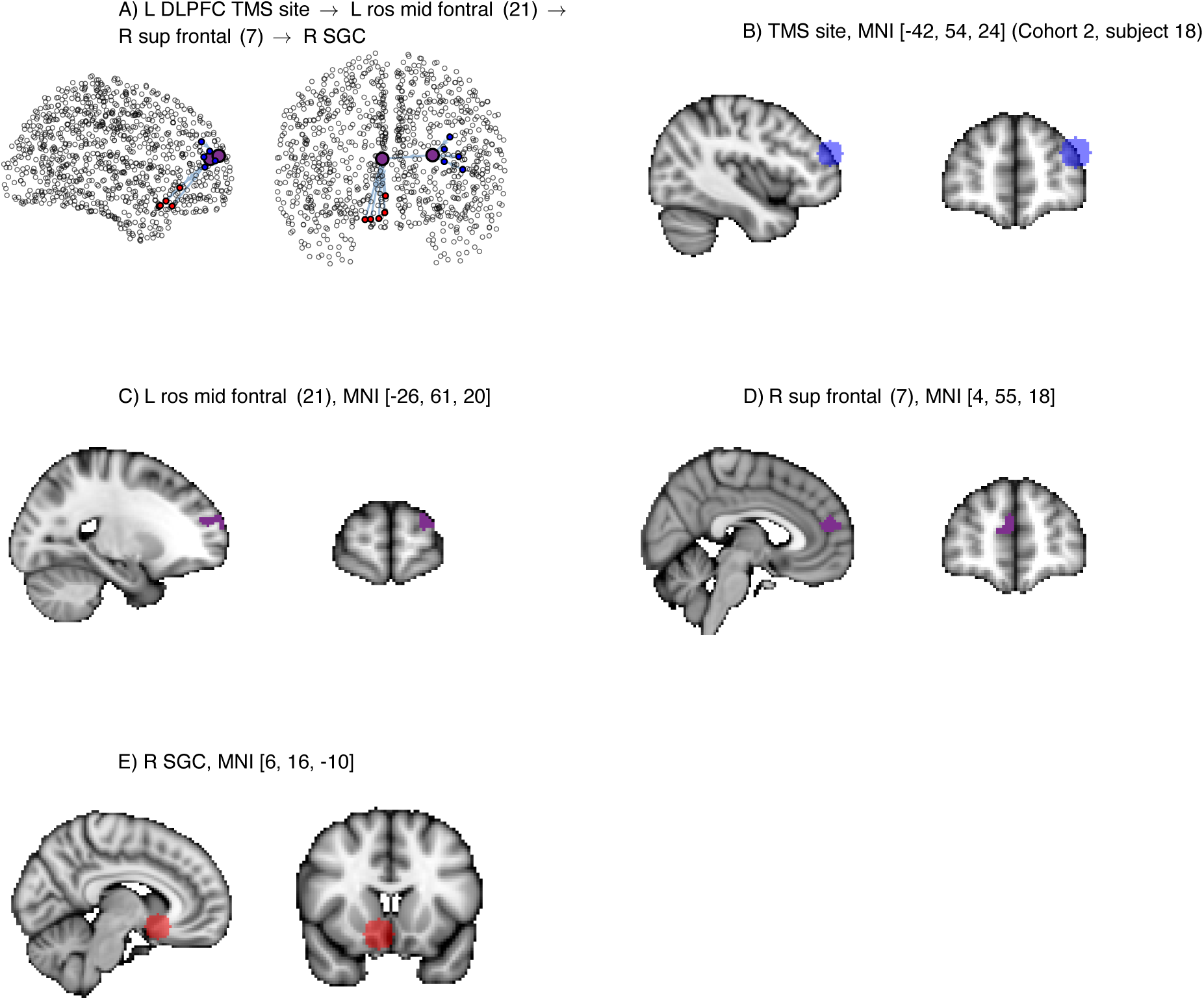
DLPFC-SGC path #1, Cohort II, visualised in network format and volumetric MNI-space for an exemplary patient. Grey matter regions along the path are identified by their Lausanne parcellation label (scale 5) and centroid MNI coordinate.

**Figure S12.**
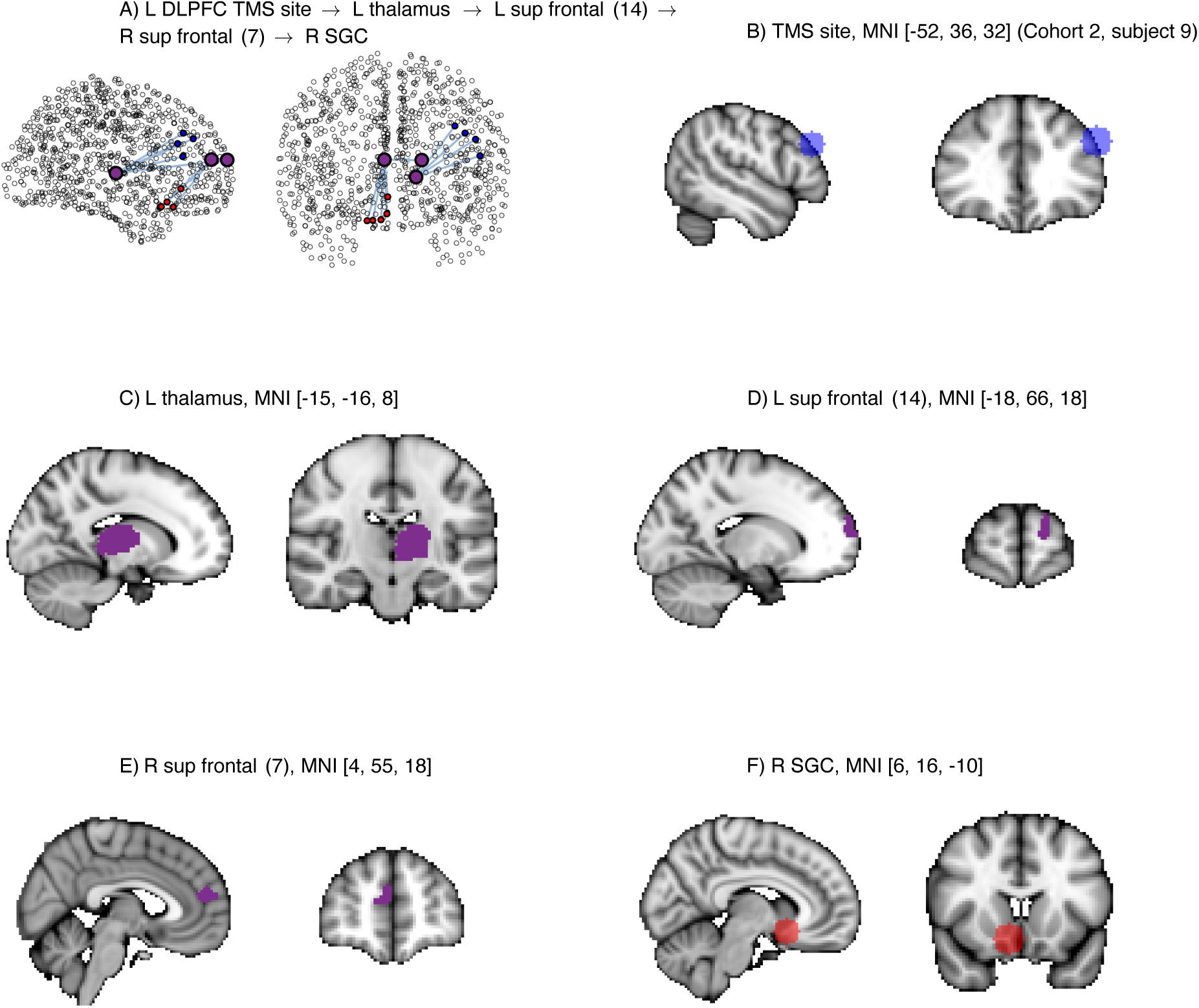
DLPFC-SGC path #2, Cohort II, visualised in network format and volumetric MNI-space for an exemplary patient. Grey matter regions along the path are identified by their Lausanne parcellation label (scale 5) and centroid MNI coordinate.

**Figure S13.**
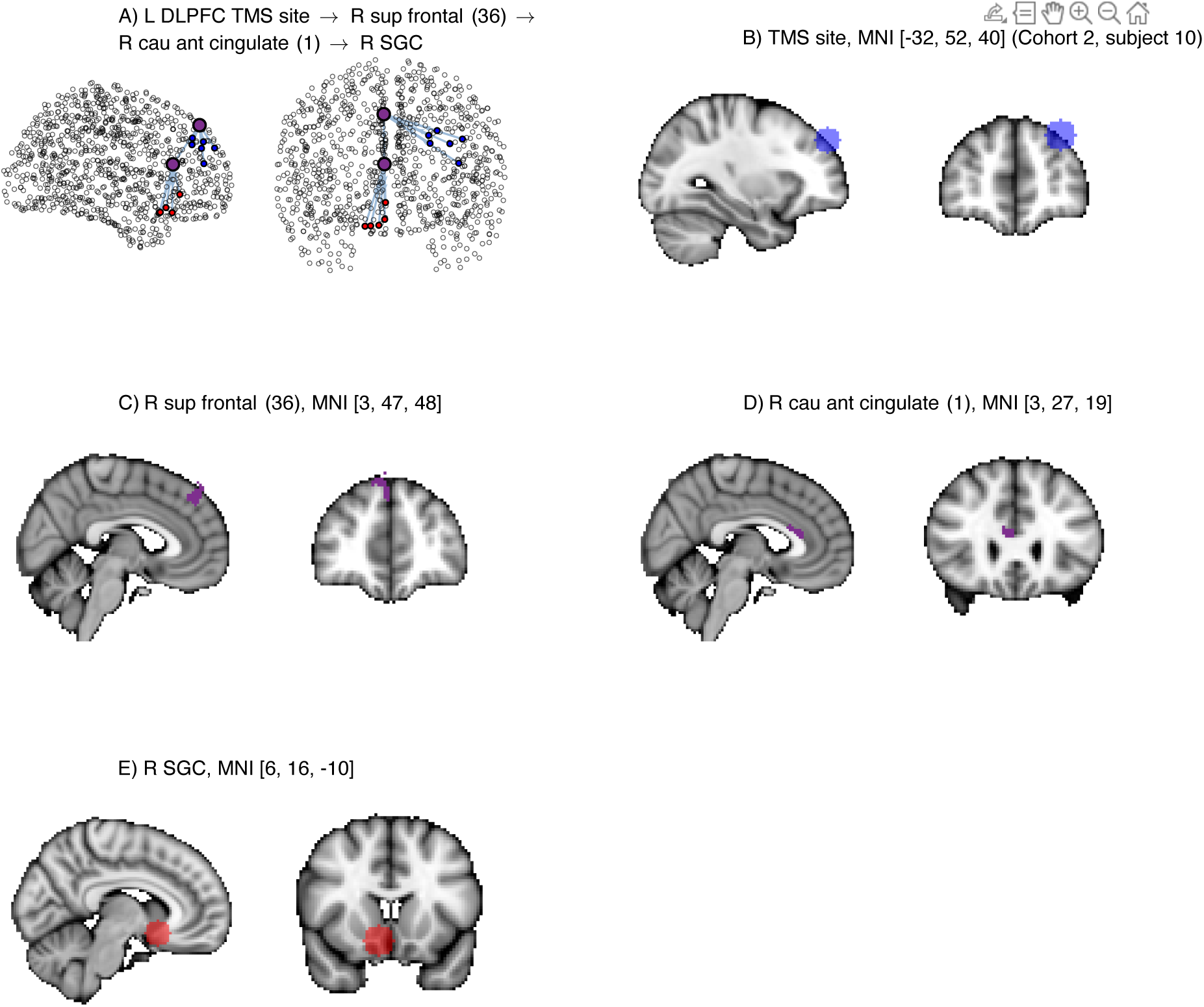
DLPFC-SGC path #3, Cohort II, visualised in network format and volumetric MNI-space for an exemplary patient. Grey matter regions along the path are identified by their Lausanne parcellation label (scale 5) and centroid MNI coordinate.

**Figure S14.**
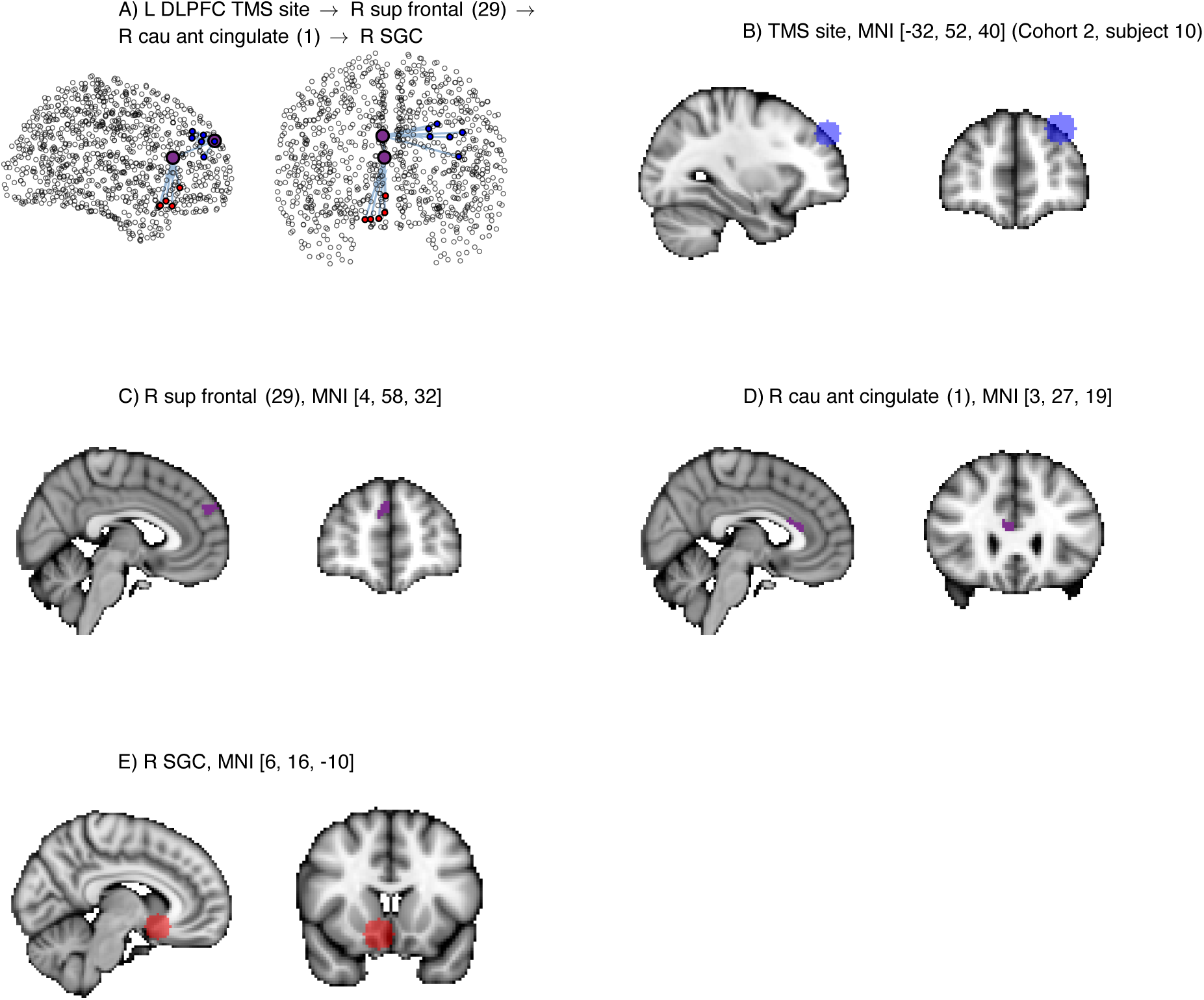
DLPFC-SGC path #4, Cohort II, visualised in network format and volumetric MNI-space for an exemplary patient. Grey matter regions along the path are identified by their Lausanne parcellation label (scale 5) and centroid MNI coordinate.

**Figure S15.**
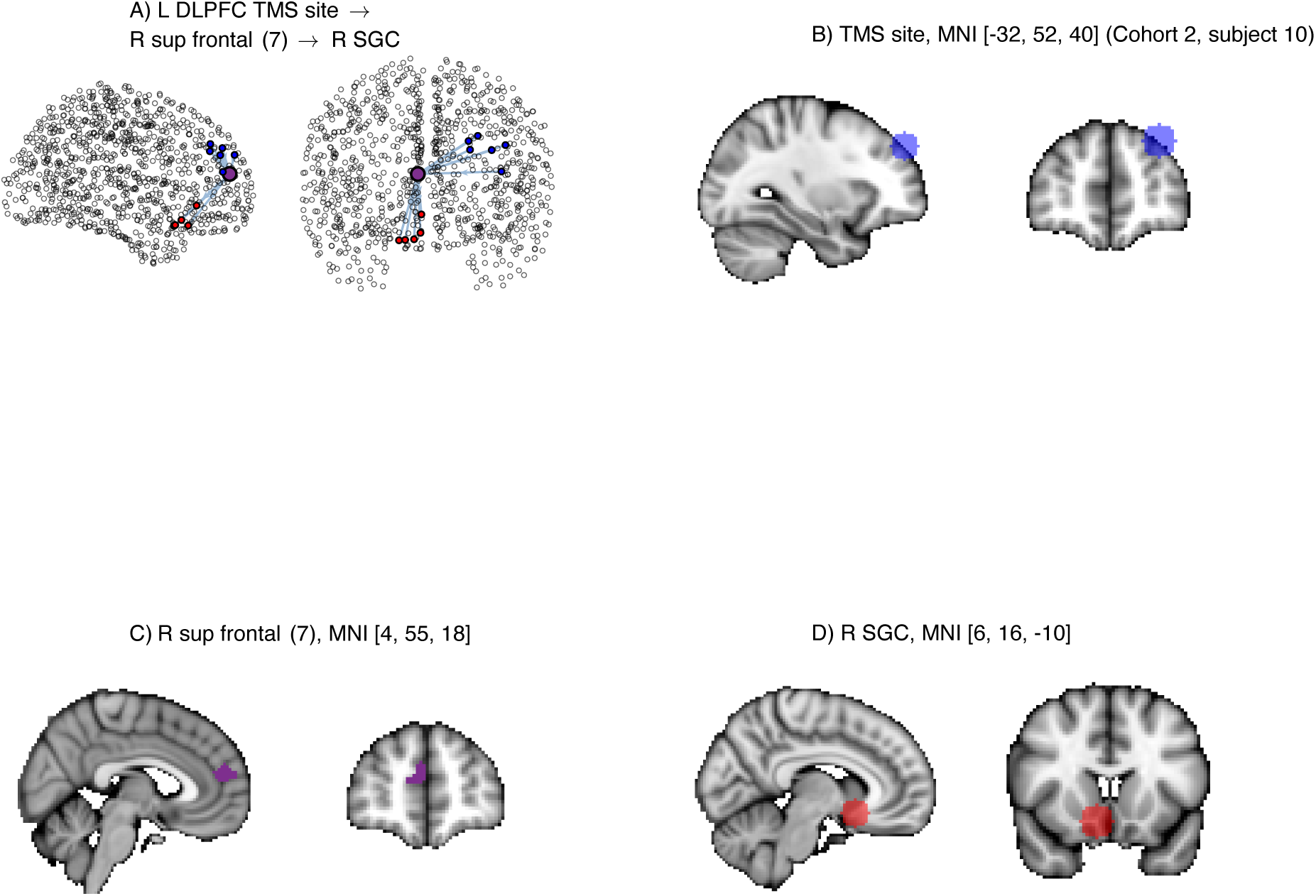
DLPFC-SGC path #5, Cohort II, visualised in network format and volumetric MNI-space for an exemplary patient. Grey matter regions along the path are identified by their Lausanne parcellation label (scale 5) and centroid MNI coordinate.

**Figure S16.**
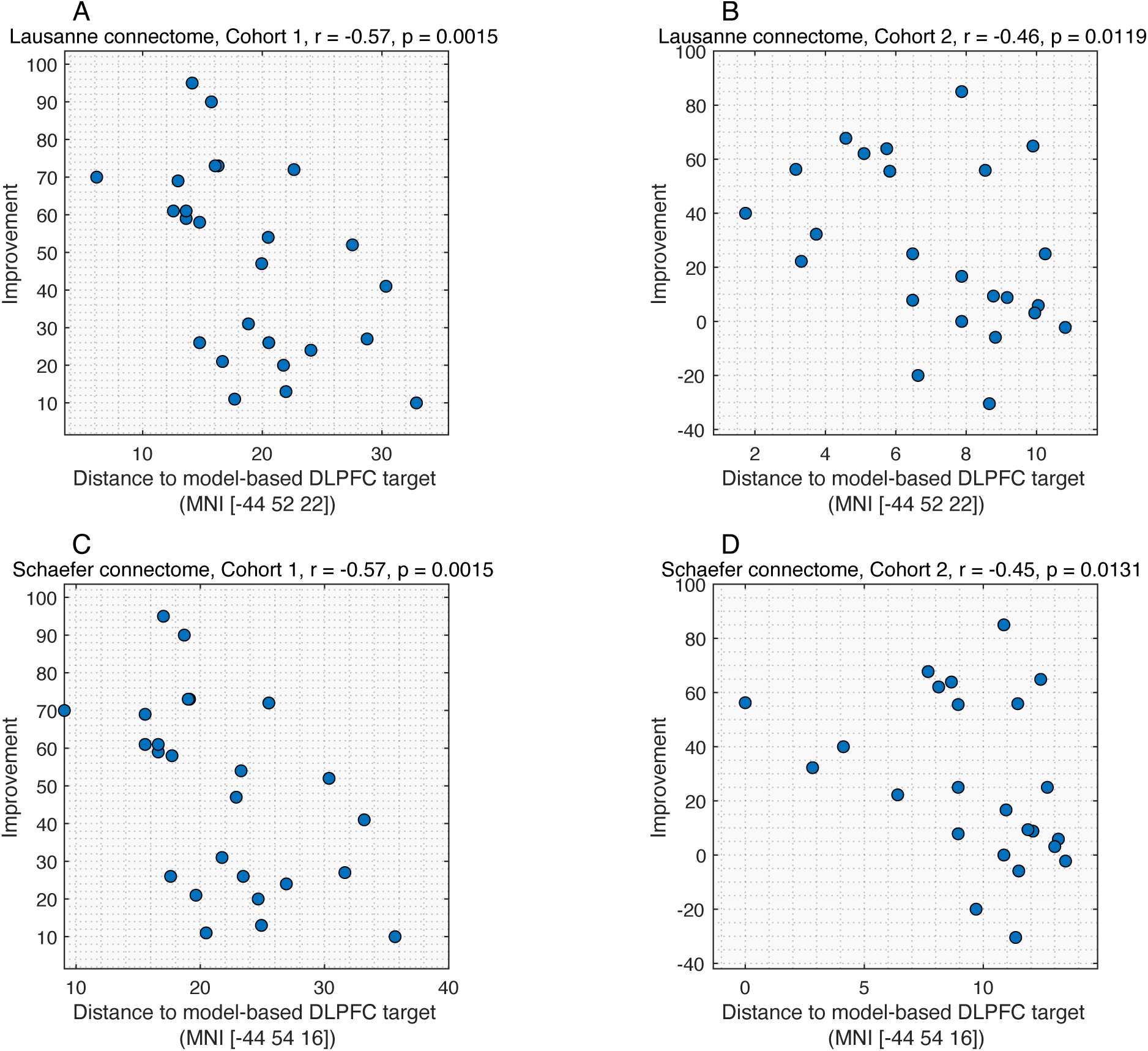
Evaluation of model-based stimulation targets. We determined the DLPFC coordinates that minimise the polysynaptic path length to the SGC (Fig 3A,D; Lausanne connectome: MNI [–44,52,22], Schaefer connectome: MNI [–44,54,16]) and computed their Euclidean distance to each patient’s stimulation site. Scatter plots show the association between treatment response and the distance to the model-based coordinates. Across both cohorts and normative connectomes, patients stimulated nearer to the DLPFC loci of minimum polysynaptic path length to the SGC showed better treatment outcomes.

**Figure S17.**
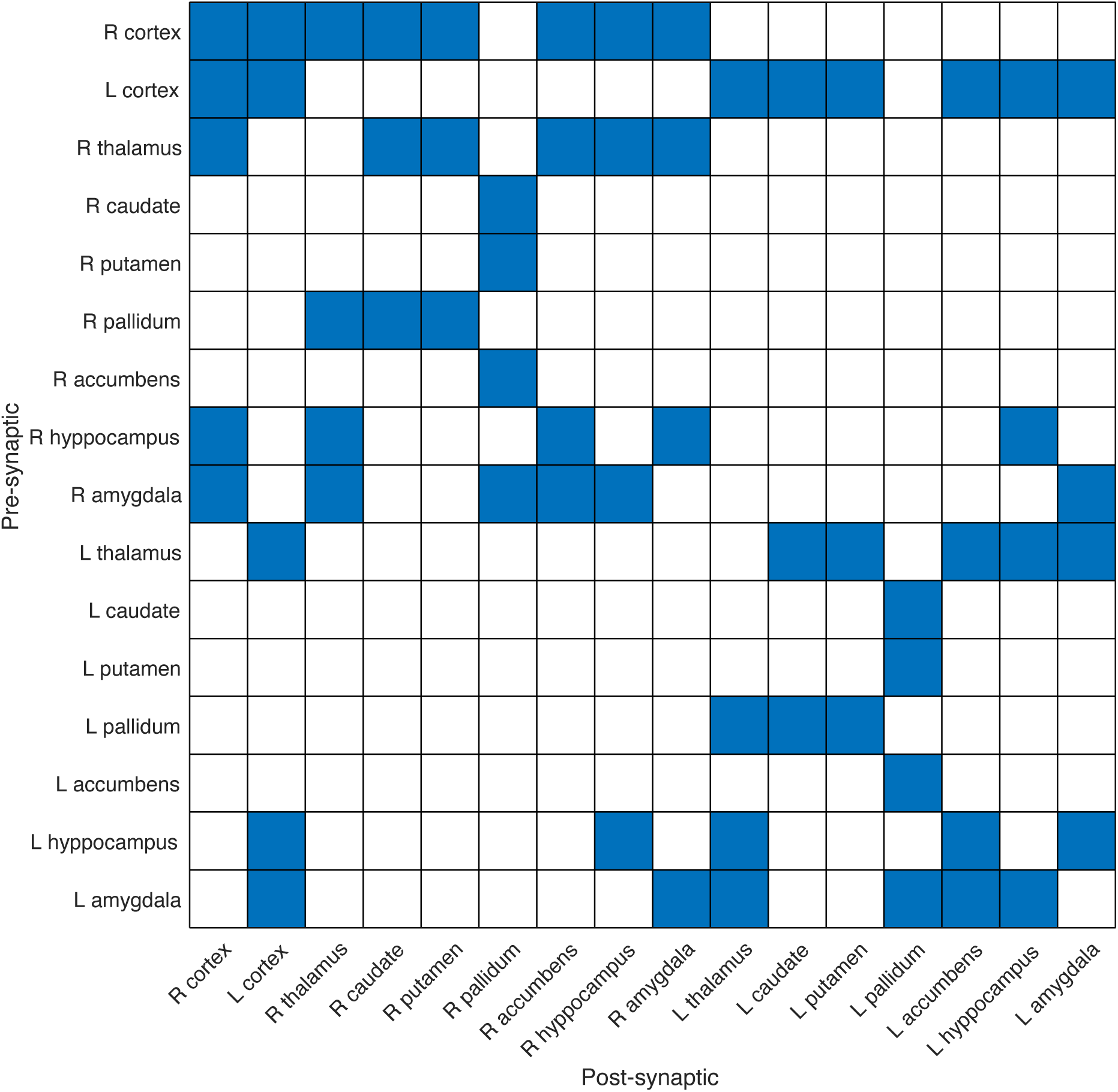
Neuroanatomical constraints used to refine structural connectivity matrices. A matrix entry *ij* indicates the presence (blue) or absence (white) of monosynaptic projections from structure *i* to structure *j*, based on primate tract-tracing studies (see *Neuroanatomically constrained connectomes*).

**Figure S18.**
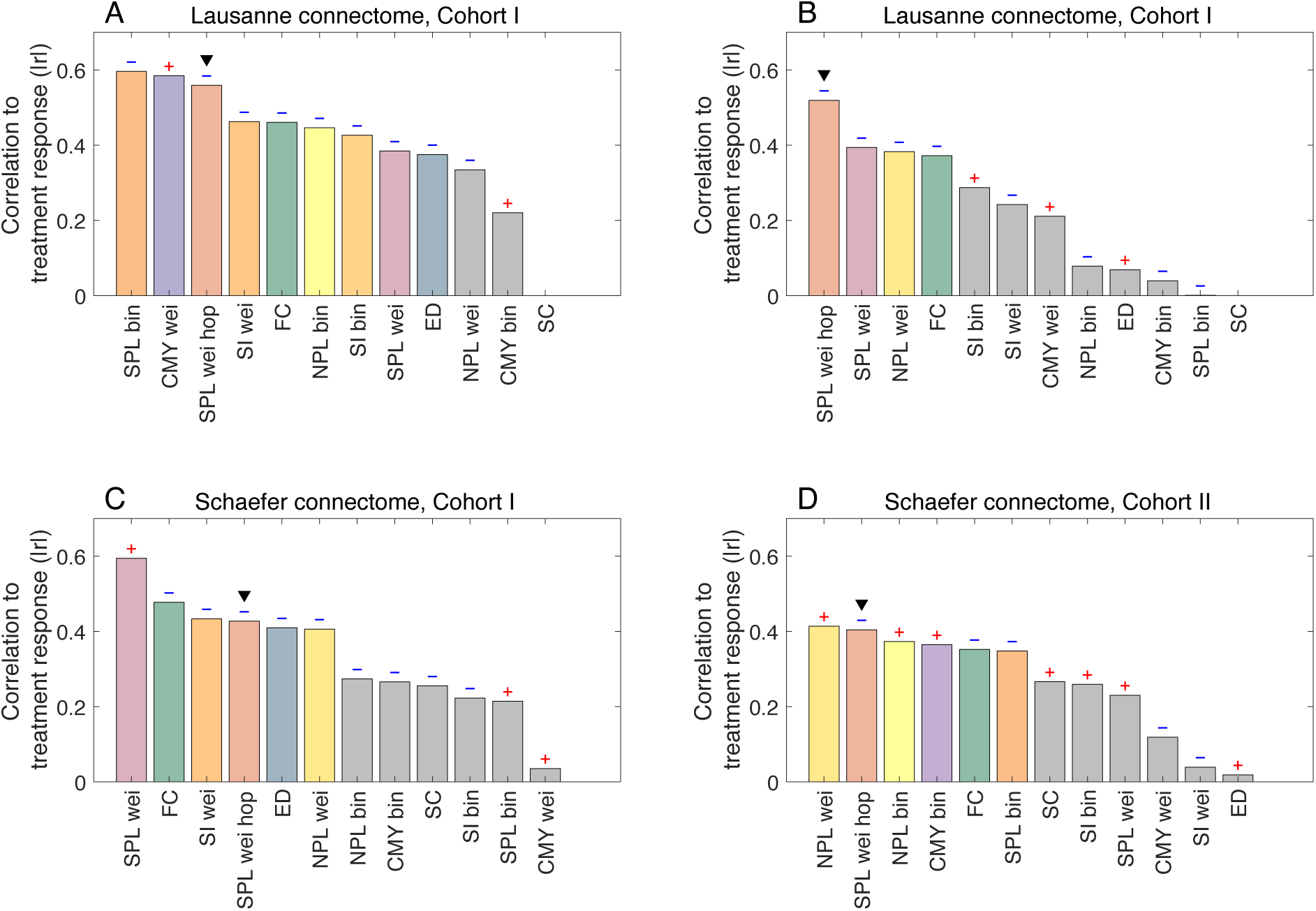
Correlations between treatment response and various connectivity, communication and distance measures, obtained using unconstrained normative connectomes. Replication of Fig S3 without performing the pre-processing step described in *Neuroanatomically constrained connectomes*.

**Figure S19.**
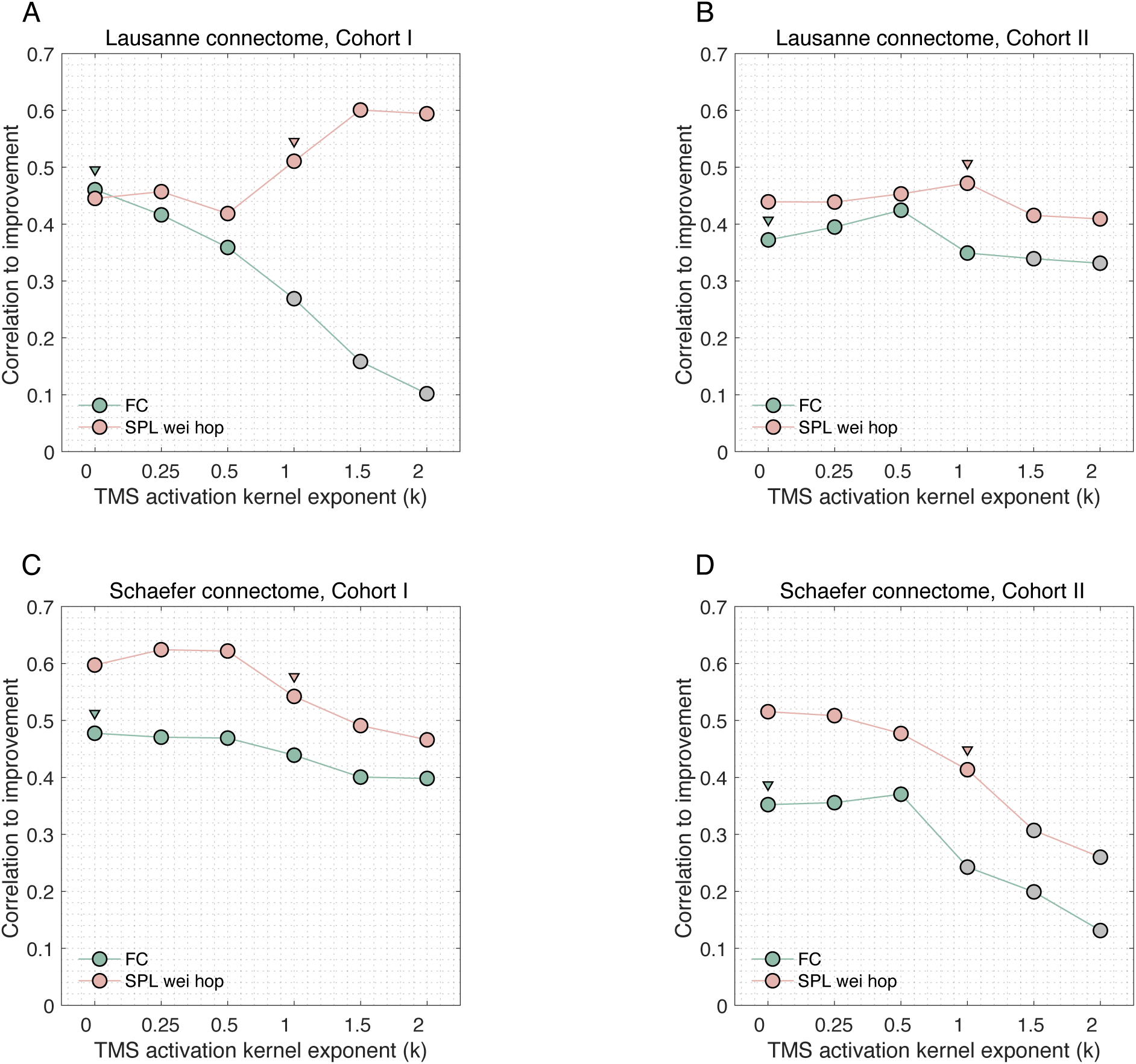
Sensitivity analyses exploring the impact of the TMS activation kernel exponent. Plots show the correlation between treatment response and DLPFC-SGC white matter hops (pink) and FC (green) remains robust for a range of *κ*. Associations that were not significant (*p* > 0.05) are marked with grey circles. For network measures, we set the decay parameter *κ* = –1 (pink triangular markers), reflecting the spatial specificity of white matter pathways, where small variations in the stimulation site may engage different discrete fibres. For functional connectivity, consistent with previous work [6–8], we used a uniform weighting (*κ* = 0; green triangular markers), which better reflects the spatial smoothness of fMRI data.

